# *Ex vivo* susceptibility to antimalarial drugs and polymorphisms in drug resistance genes of African *Plasmodium falciparum*, 2016-2023: a genotype-phenotype association study

**DOI:** 10.1101/2024.07.17.24310448

**Authors:** Jason Rosado, Abebe A. Fola, Sandrine Cojean, Véronique Sarrasin, Romain Coppée, Rizwana Zaffaroulah, Azza Bouzayene, Liliane Cicéron, Ludivine Houzé, Rebecca Crudale, Lise Musset, Marc Thellier, Bruno Pradines, Jérôme Clain, Jeffrey A. Bailey, Sandrine Houzé, Investigation Study Group

## Abstract

**Background:** Given the altered responses to both artemisinins and lumefantrine in Eastern Africa, monitoring antimalarial drug resistance in all African countries is paramount.

**Methods:** We measured the susceptibility to six antimalarials using *ex vivo* growth inhibition assays (IC_50_) for a total of 805 *Plasmodium falciparum* isolates obtained from travelers returning to France (2016-2023), mainly from West and Central Africa. Isolates were sequenced using molecular inversion probes (MIPs) targeting fourteen drug resistance genes across the parasite genome.

**Findings:** *Ex vivo* susceptibility to several drugs has significantly decreased in 2019-2023 versus 2016-2018 parasite samples: lumefantrine (median IC_50_: 23·0 nM [IQR: 14·4-35·1] in 2019-2023 versus 13·9 nM [8·42-21·7] in 2016-2018, p<0·0001), monodesethylamodiaquine (35·4 [21·2-51·1] versus 20·3 nM [15·4-33·1], p<0·0001), and marginally piperaquine (20·5 [16·5-26·2] versus 18.0 [14·2-22·4] nM, p<0·0001). Only four isolates carried a validated *pfkelch13* mutation. Multiple mutations in *pfcrt* and one in *pfmdr1* (N86Y) were significantly associated with altered susceptibility to multiple drugs. The susceptibility to lumefantrine was altered by *pfcrt* and *pfmdr1* mutations in an additive manner, with the wild-type haplotype (*pfcrt* K76-*pfmdr1* N86) exhibiting the least susceptibility.

**Interpretation:** Our study on *P. falciparum* isolates from West and Central Africa indicates a low prevalence of molecular markers of artemisinin resistance but a significant decrease in susceptibility to the partner drugs that have been the most widely used since a decade –lumefantrine and amodiaquine. These phenotypic changes likely mark parasite adaptation to sustained drug pressure and call for intensifying the monitoring of antimalarial drug resistance in Africa.

**Funding:** This work was supported by the French Ministry of Health (grant to the French National Malaria Reference Center) and by the Agence Nationale de la Recherche (ANR-17-CE15-0013-03 to JC). JAB was supported by NIH R01AI139520. JR postdoctoral fellowship was funded by Institut de Recherche pour le Développement.

**Research in context:** *Evidence before this study:* Artemisinin-based combination therapies (ACTs) have been introduced since the 2000s as the first-line curative treatment of malaria. ACTs combine an artemisinin derivative, which rapidly reduces parasite load, with another antimalarial drug –known as partner drug-which eliminates the remaining parasites thanks to its longer half-life. This approach reduces the likelihood of parasites developing resistance to both drugs, thereby increasing treatment efficacy and delaying the emergence of resistance. However, resistance to artemisinins and then to some partner drugs was identified in Southeast Asia more than a decade ago and has spread throughout the region. Artemisinin partial resistance is now emerging in the East and Horn of Africa. It manifests as delayed parasite clearance from the bloodstream after treatment, increasing the parasite load in contact with the partner drug only and the likelihood of selecting resistant parasites. It is, therefore, important to monitor antimalarial drug susceptibility and drug resistance mutations in contemporary African isolates, especially in the understudied West and Central African regions, to anticipate the spread of multidrug-resistant parasites. We searched for articles on antimalarial drug resistance published between January 1, 2000, and July 1, 2024, using the PubMed search terms “antimalarial resistance”, “Africa”, and “*ex vivo*”. Of the 69 published studies, only six encompassing a total of 827 isolates across five West and Central African countries from 2016 to 2022 combined *ex vivo* drug assays with genotyping data. Parasites with an increased rate of *ex vivo* survival to artemisinins were reported in one study from Ghana (7/90 isolates in 2018) and another from The Gambia (4/41 isolates in 2017). Only the Ghanaian study reported mutations in the non-propeller domain of *pfkelch13* gene, whereas the Gambian study reported mutations associated with reduced susceptibility to lumefantrine (7%, 3/41). In Mali, Senegal and Burkina Faso, most isolates were susceptible to commonly used antimalarial drugs (chloroquine, amodiaquine, piperaquine, mefloquine, lumefantrine and dihydroartemisinin) using standard growth inhibition assays. In Ghana, reduced susceptibility to artemisinin, mefloquine and amodiaquine was observed. The relative lack of recent data on parasite susceptibility to antimalarial drugs in recent parasites from West and Central Africa prompted us to conduct this study.

*Added value of this study:* *Ex vivo* susceptibility to six antimalarial drugs (dihydroartemisinin, lumefantrine, mefloquine, chloroquine, monodesethylamodiaquine, and piperaquine) and mutations in fourteen drug resistance genes were evaluated in 805 isolates collected between January 2016 and February 2023 from 35 African countries, mainly from West and Central Africa. Median IC_50_ values were in the low nanomolar range, indicating good potency against *P. falciparum*. However, worrying trends emerged from 2019 onwards, with median IC_50_ values for lumefantrine that increased from 13·9 nM in 2016-18 to 23 nM in 2019-23 and for amodiaquine from 20·3 nM to 35·4 nM. The high prevalence of resistance alleles in *pfdhfr*, *pfdhps*, *pfmdr1* and *pfcrt* genes underscores the sustained pressure exerted by antimalarial drugs on parasite populations. Notably, although the triple mutant *pfdhfr* N51I-C59R-S108N was highly prevalent, the *dhfr-dhps* quintuple mutant (with extra *pfdhps* A437G-K540E), which is responsible for sulfadoxine-pyrimethamine treatment failure in adults and children, was rare. In addition, the analysis revealed some geographic and temporal variations in mutation prevalence. The genotype-phenotype association analysis performed in this study elucidates the relationship between genetic variants and *ex vivo* drug susceptibility, providing valuable information for understanding the molecular basis of resistance and informing future treatment strategies. For example, mutations in the *pfcrt* and *pfmdr1* genes, mainly K76T and N86Y, were associated with altered susceptibilities to most drugs. Haplotypic association analysis further indicated that the two genes have cumulative effects on the susceptibility to lumefantrine, with the wild-type haplotype (*pfcrt* K76-*pfmdr1* N86) exhibiting the least susceptibility.

*Implication of all the available evidence:* While the susceptibility to most antimalarials suggests continued efficacy, the observed decrease in susceptibilities to lumefantrine and amodiaquine in parasites from West and Central Africa from 2019 onwards suggests an ongoing adaptation of parasites, possibly related to the increasing use of ACT treatments in Sub-saharan Africa since a decade. These phenotypic changes over time were accompanied by small changes in the prevalence of resistance alleles in *pfcrt* and *pfmdr1* genes. Additional changes, potentially leading to larger decreases in drug susceptibilities, can be expected over time. The large-scale analysis presented here provides invaluable, contemporary insights into the current landscape of susceptibility to antimalarial drugs and molecular markers of resistance in *P. falciparum* isolates from West and Central Africa. While the data suggests that ACTs and sulfadoxine-pyrimethamine are likely to be effective in these regions, the phenotypic changes we observed call for intensifying the monitoring of antimalarial drug resistance in Africa.

## Introduction

Malaria continues to take a heavy toll on public health in Africa. Antimalarials remain a cornerstone in the fight against malaria, and the evolution of drug resistance poses a significant threat to the efficacy of treatment regimens.^1,2^ Monitoring antimalarial drug resistance and treatment efficacy is paramount, as drug selection pressures are ongoing, and no alternatives to current first-line treatments will be available in the coming years.^3,4^ Artemisinin-based combination therapies (ACTs) as first-line curative treatments that were introduced in the 2000s. ACTs combine an artemisinin derivative, which rapidly reduces parasite load, with another antimalarial drug –known as partner drug-which eliminates the remaining parasites thanks to its longer half-life.

However, the emergence of artemisinin partial resistance (ART-R) has raised concerns about the long-term efficacy of ACTs for effective malaria treatment.^5,6^ After ART-R emerged, treatment failures with dihydroartemisinin (DHA)-piperaquine (PPQ) were reported in Southeast Asia as parasites acquired additional resistance to PPQ.^7,8^ ART-R has recently emerged in East Africa and the Horn of Africa,^9–11^ creating the potential for selection of parasites resistant to the partner drug(s) and threatening the long-term efficacy of ACTs in Africa.^12,13^ Worryingly, decreased susceptibility of *P. falciparum* isolates to the partner drug lumefantrine (LMF) has also been reported in northern Uganda.^14,15^ In West and Central Africa, treatment efficacy studies have confirmed the clinical efficacy of ACTs in several settings overall.^16,17^ However, few studies have reported *ex vivo* antimalarial drug assays for West and Central African isolates, with little evidence of reduced susceptibility to partner drugs.^18,19^ Molecular surveillance has also revealed the sporadic presence of validated ART-R *pfkelch13* mutations in these two regions.^20^

To better understand the current situation in West and Central Africa, we analyzed the *ex vivo* susceptibility to six drugs (DHA, LMF, mefloquine (MFQ), chloroquine (CQ), monodesethylamodiaquine (MDAQ), and PPQ; assessed by growth inhibition assays) of 805 *P. falciparum* isolates collected between 2016 and 2023 from imported malaria cases in returned travelers to mainly West and Central Africa. The isolates were also sequenced using high-throughput molecular inversion probes (MIPs) targeting 815 overlapping targets across fourteen drug resistance genes to determine the prevalence of drug resistance markers and to estimate genetic correlates of drug susceptibility phenotypes.

## Methods

### Sample and data collection

The French National Malaria Reference Center (FNMRC) coordinates a nationwide network of hospitals in mainland France to survey imported malaria. Blood samples from civilian or military hospitals participating in the FNRMC network were sent to the FNRMC reference laboratory (Bichat Hospital in Paris) (see Study Group). Inclusion criteria were: diagnosis of *P. falciparum* malaria recorded between January 2016 and February 2023 and confirmed by microscopy and PCR, available DNA sample, information on the country visited, and available IC_50_ data against CQ, MFQ, PPQ, MDAQ, LMF and DHA. A total of 813 isolates fulfilled the inclusion criteria.

### Ex vivo assays

CQ, DHA, MDAQ, MFQ, LMF, and PPQ were purchased from Alsachim (Illkirch Graffenstaden, France). The susceptibility of the isolates to the antimalarial drugs was assessed without culture adaptation. 100 µL of parasitized erythrocytes (final parasitaemia of 0.5% and a final hematocrit of 1.5%) were aliquoted into 96-well plates preloaded with a concentration gradient of antimalarial drugs. The plates were incubated for 48 hours in a controlled atmosphere of 85% N_2_, 10% O_2_, 5% CO_2_ at 37°C. Drug susceptibilities were determined as half-maximal inhibitory concentrations (IC_50_s) using the standard 42-hour [^3^H]hypoxanthine uptake inhibition assay (**Supplementary Methods**) ^21^. Resistance thresholds for the six compounds were previously reported by Pradines, Kaddouri and others.^22–24^

### DNA extraction and MIP sequencing of drug resistance genes

Genomic DNA was extracted from 200 μL whole blood using a MagNA Pure automaton (Roche Diagnostics, USA), eluted in 100 μL, and stored at –20°C. DNA extracts from the included samples were sent to Brown University for MIP sequencing. All samples were then genotyped using molecular inversion probes (MIPs, n=815) targeting known drug resistance SNP mutations in 14 genes across the *P. falciparum* genome, as previously described (**Table S1, Appendix 2)**.^12,25^ Sequencing was performed on an Illumina NextSeq 550 instrument (150 bp paired-end reads) at Brown University (RI, USA).

### MIP data analysis and estimating drug resistance prevalence

Sequencing data processing and variant calling were performed using MIPtools (v0.19.12.13; https://github.com/bailey-lab/MIPTools). The prevalence for each drug resistance marker was calculated as p = x / n * 100, where p is the prevalence, x is the number of infections with mutant alleles, and n is the number of successfully genotyped infections, as previously described.^12^ Mixed genotypes, i.e. a sample with a reference allele and an alternative allele in a given sample, were considered mutant regardless of the within-sample mutant allele frequency. Haplotypes were reconstructed for all samples using the major allele in a given codon (**Supplementary Methods**).

### Phenotype–genotype association analysis

Isolates with >30% missing genotype data were discarded. We then retained only biallelic non-synonymous SNPs with minor allele frequency (MAF) >0.01 for phenotype-genotype association analysis. Phenotype-genotype association was performed with the SNPassoc R package (v.2.1.0) ^26^ using the built-in WGassociation function under a dominant model. For each drug, a linear regression model was fitted with continuous IC_50_ values as the outcome and individual SNP genotypes as independent variables. The model was adjusted for the complexity of infection (COI), year, and the visited country of imported cases as covariates.

### Ethical aspects

Informed consent was not required because the clinical and biological data were collected from the FNMRC database by the common public health mission of all National Reference Centers in France, in coordination with the Santé Publique France organization for malaria surveillance and care. According to article L1221–1.1 of the Public Health Code in France, the study was considered non-interventional research and only required the patient’s non-opposition (according to article L1211–2 of the Public Health Code). All data were anonymized before use. The FNMRC got the authorization to use their databases according to the National Data Protection Commission (CNIL) (number 1223103). Human DNA was not analyzed.

## Results

### Geographical origin and year of collection

From January 2016 to February 2023, 1,126 *P. falciparum* isolates from travelers were evaluated for *ex vivo* antimalarial drug susceptibility at the FNMRC laboratory. Of these, 813 isolates were included in this study, corresponding to samples with validated IC_50_ for the following six drugs: DHA, LMF, MFQ, MDAQ, CQ, and PPQ. According to reported travel history, infections originated from thirty-five African countries, but most isolates were from travelers who had visited West (59%, 471/805) and Central (38%, 306/805; **Figure 1A**) African countries. The most visited countries were Côte d’Ivoire (27%, 217/805), Cameroon (21%, 169/805), Mali (9%, 69/805), Guinea (7%, 58/805), and Republic of the Congo (6%, 45/805) (**Figure 1A** and **Table S2**). The number of samples collected per year was higher in 2016 (165/805), 2017 (166/805) and 2018 (167/805) than in 2019 (100/805) and beyond (**Figure 1B**). No differences were seen in the distribution of countries between 2016-2018 and 2019-2023 (p>0·05, **Table S3**). Travelers were predominantly male (62%), with a median age of 40 years and a median parasitemia of 1·7% (**Table 1**).

**Figure 1.**
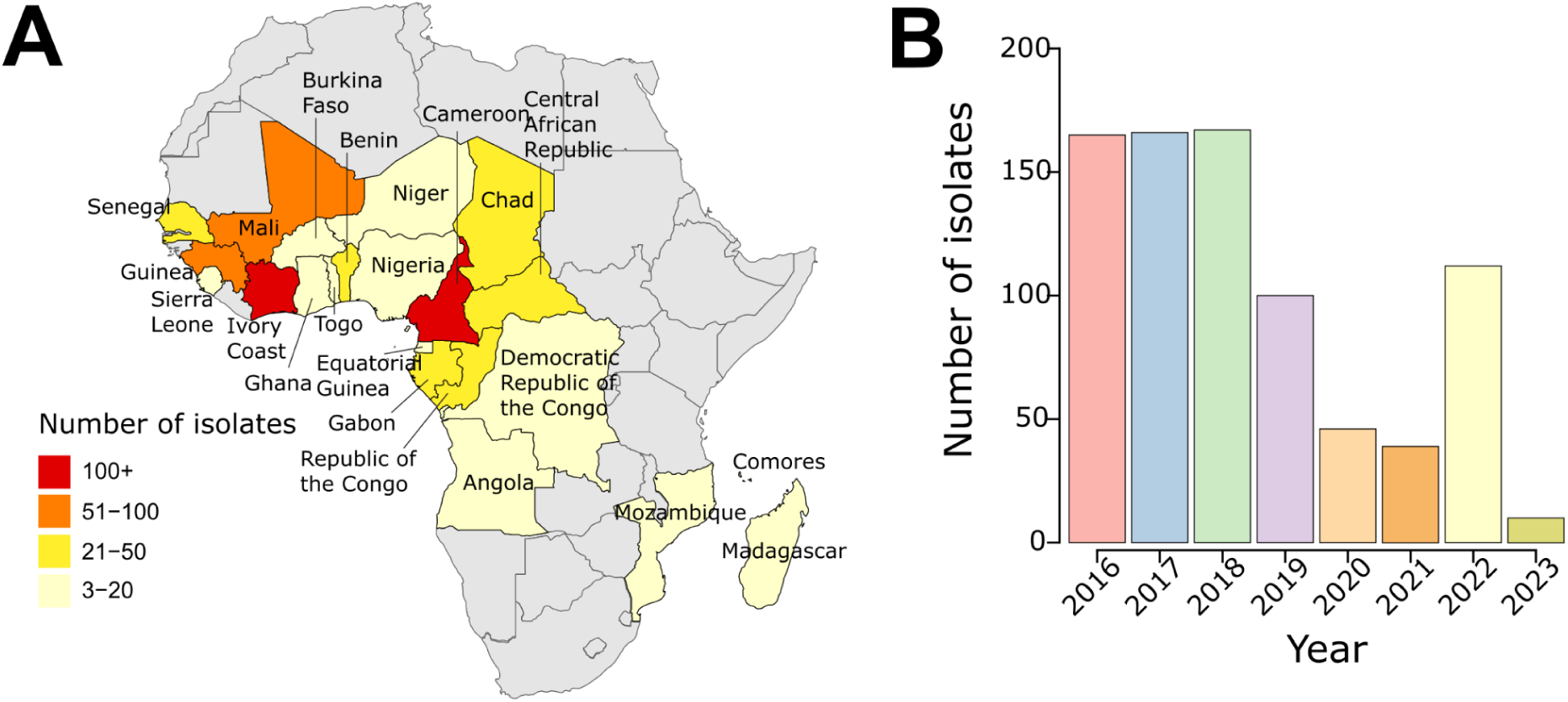
Geographical origin and collection year of isolates. **A**) Map of Africa showing the origin of malaria cases imported into France. Colors indicate sample size, and countries with fewer than three isolates are not shown. **B)** Bar plot showing the temporal distribution of imported malaria cases included in this study.

**Table 1.**
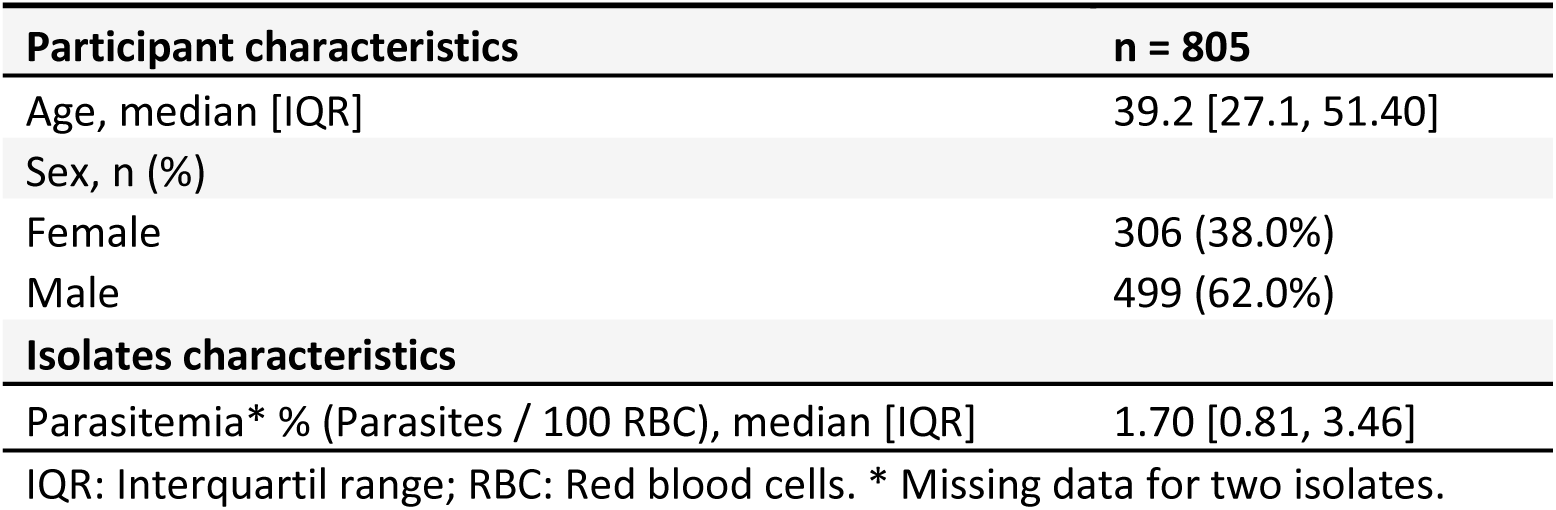
Epidemiological characteristics of travelers returning to France and description of isolates from 2016 – 2023.

| Participant characteristics | n = 805 |
| --- | --- |
| Age, median [IQR] | 39.2 [27.1, 51.40] |
| Sex, n (%) |  |
| Female | 306 (38.0%) |
| Male | 499 (62.0%) |
| Isolates characteristics |  |
| Parasitemia* % (Parasites / 100 RBC), median [IQR] | 1.70 [0.81, 3.46] |

IQR: Interquartil range; RBC: Red blood cells. \* Missing data for two isolates.

### Half-maximal inhibitory concentration (IC_50_) for six antimalarial compounds

IC_50_ values of *P falciparum* 3D7 laboratory reference strain are indicated in **Table S4**. For the isolates tested in this study, the median IC_50_ estimated with the standard growth inhibition assay was less than 30 nM for all assessed antimalarial compounds, consistent with potent activity. The median IC_50_ values for the six drugs were 1·1 nM [IQR: 0·8-1·7] for DHA, 16·7 nM [9·9-27·4] for LMF, 29·5 nM [19·1-45·5] for MFQ, 23·4 nM [17·1-39·0] for MDAQ, 26·7 nM [18·0-41·2] for CQ, and 18·5 nM [15·1-24·3] for PPQ. Using published resistance thresholds,^22–24^ 49% of the isolates were classified as resistant to MFQ (IC_50_ > 30 nM), 10% to CQ (IC_50_ > 100 nM), and 3% to MDAQ (IC_50_ > 80 nM). Less than 1% of isolates had an IC_50_ above the threshold for DHA (10 nM), LMF (150 nM) and PPQ (135 nM) (**Figure S1**).

Remarkably, when compared to samples collected in 2016-2018, a significant decrease in susceptibility was observed in 2019-2023 samples for LMF (median IC_50_: 13·9 nM [IQR: 8·42-21·7] in 2016-2018 versus 23·0 nM [14·4-35·1] in 2019-2023; p<0·001), MDAQ (20·3 nM [15·4-33·1] versus 35·4 nM [21·2-51·1]; p<0·001), and modestly for PPQ (18·0 nM [14·2-22·4] versus 20·5 [16·5-26·2]; p<0·001) (**Figure 2, Table S5**). In contrast, a modest improvement in susceptibility was observed in 2019-2023 samples for CQ (27·7 nM [19·0-45·0] versus 25·1 nM [17·3-38·9]; p=0·05), and there was no significant change for MFQ. Altered drug susceptibilities between the two time periods were again detected after data stratification into West and Central African countries, except for DHA, which was significantly altered only in samples from Central Africa, and for CQ (**Table S6**), possibly because of less power for CQ. We note that these temporal changes in drug susceptibility were not significant when using 1-year bins during the 2016-2023 study period (**Table S7**). The frequency of CQ-resistant isolates (IC_50_ > 100 nM) decreased nearly by half in 2019-2023 compared to 2016-2018 (7% in 2019-2023 versus 12% in 2016-2018, p<0·05) (**Table S8**).

**Figure 2.**
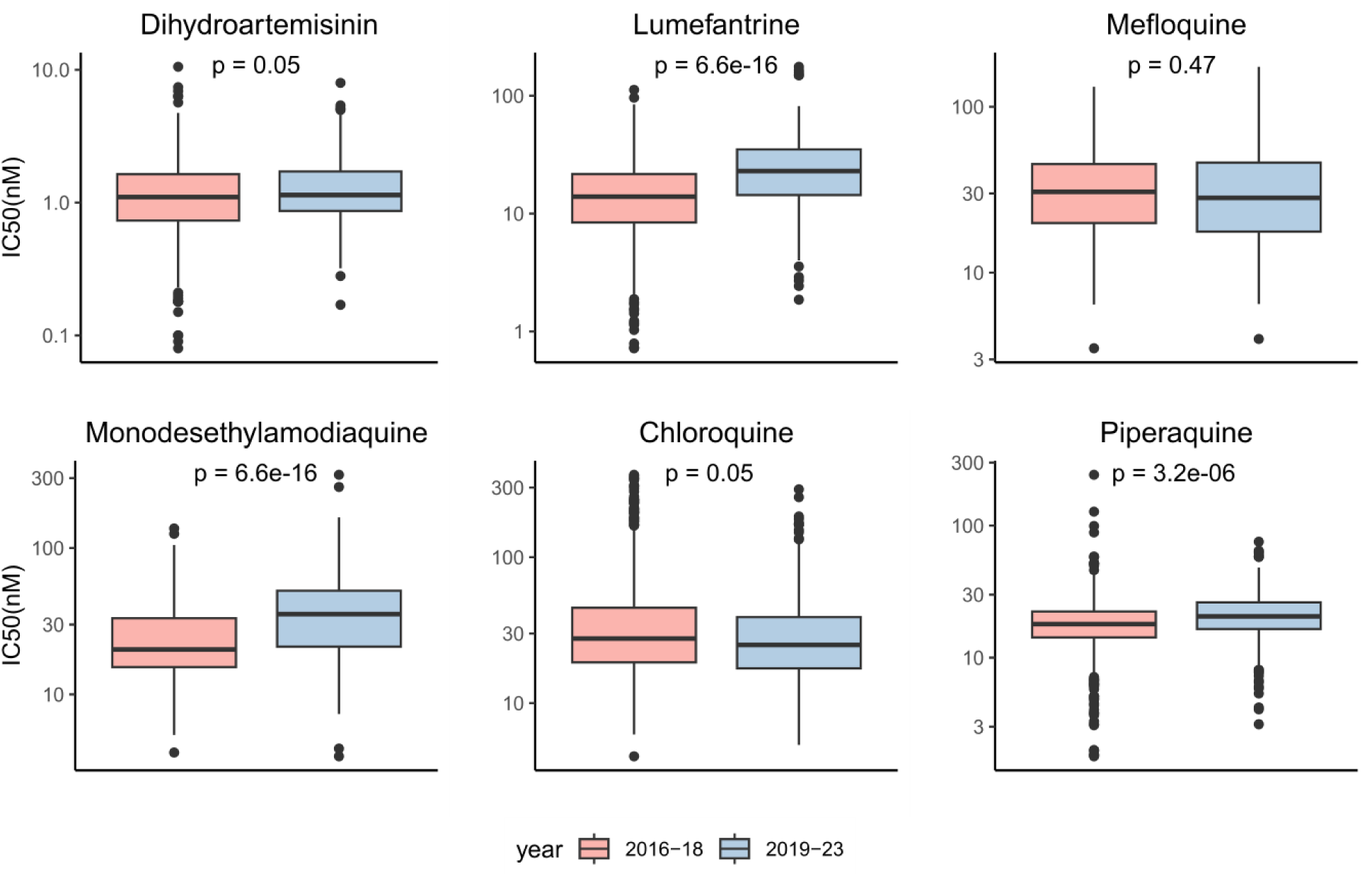
Half-maximal inhibitory concentration (IC_50_) for six antimalarial drugs. **A**) Distribution of IC_50_ for the six antimalarial drugs in 2016-18 and 2019-23. Box plots show the median IC_50_ (in nM) and interquartile range. Change in susceptibility over time was tested by Mann-Whitney test. P-values were adjusted using Benjamini-Hochberg correction.

There were several significant drug-drug IC_50_ correlations, most of which were positive (**Figure S2**). The largest positive correlations involved the following drug pairs: LMF-MFQ, CQ-MDAQ, DHA-LMF, and DHA-MFQ. The negative correlations were much less intense and involved CQ-LMF and CQ-MFQ.

### Prevalence of validated antimalarial resistance mutations

Sequence data generated with MIPs covered 43 exons from 14 genes reported to be associated with drug resistance. 805 out of the 813 included isolates were successfully genotyped. The average number of reads per sample was 120 (range: 48-258) (**Figure S3**). Nearly 60% of the isolates (477/805) had a COI >1. We first examined the prevalence of mutations in five robustly validated drug resistance genes: *pfdhfr* and *pfdhps* (associated with resistance to pyrimethamine and sulfadoxine, respectively), *pfcrt* and *pfmdr1* (associated with multidrug resistance), and *pfkelch13* (associated with ART-R).

More than 85% of isolates carried the *pfdhfr* N51**I**, C59**R**, and S108**N** mutations (**Figure 3A**), which were predominantly found as the **IRN** triple mutant haplotype and marginally as the **I**C**N** and N**RN** double mutants (**Figure S4**), regardless of the country of infection (**Figure S5, Table S9**). We found a low prevalence of the *pfdhps* I431**V** (8.5%), K540**E** (6%), A581**G** (8%) and A613**S** (15%) mutations, and a large prevalence of S436**A** and A437**G** (53% and 78%, respectively). At *pfdhps* codons 436-437-540-581 considered jointly (haplotypes, **Figure S4**), single mutants **A**AKA and S**G**KA were the most common haplotypes (35% and 50% respectively). The quintuple mutant *dhfr+dhps* haplotype **IRN**+A437**G**-K540**E**, which is responsible for SP treatment failure in adults and children, was detected in 3% of samples (19/600).

**Figure 3.**
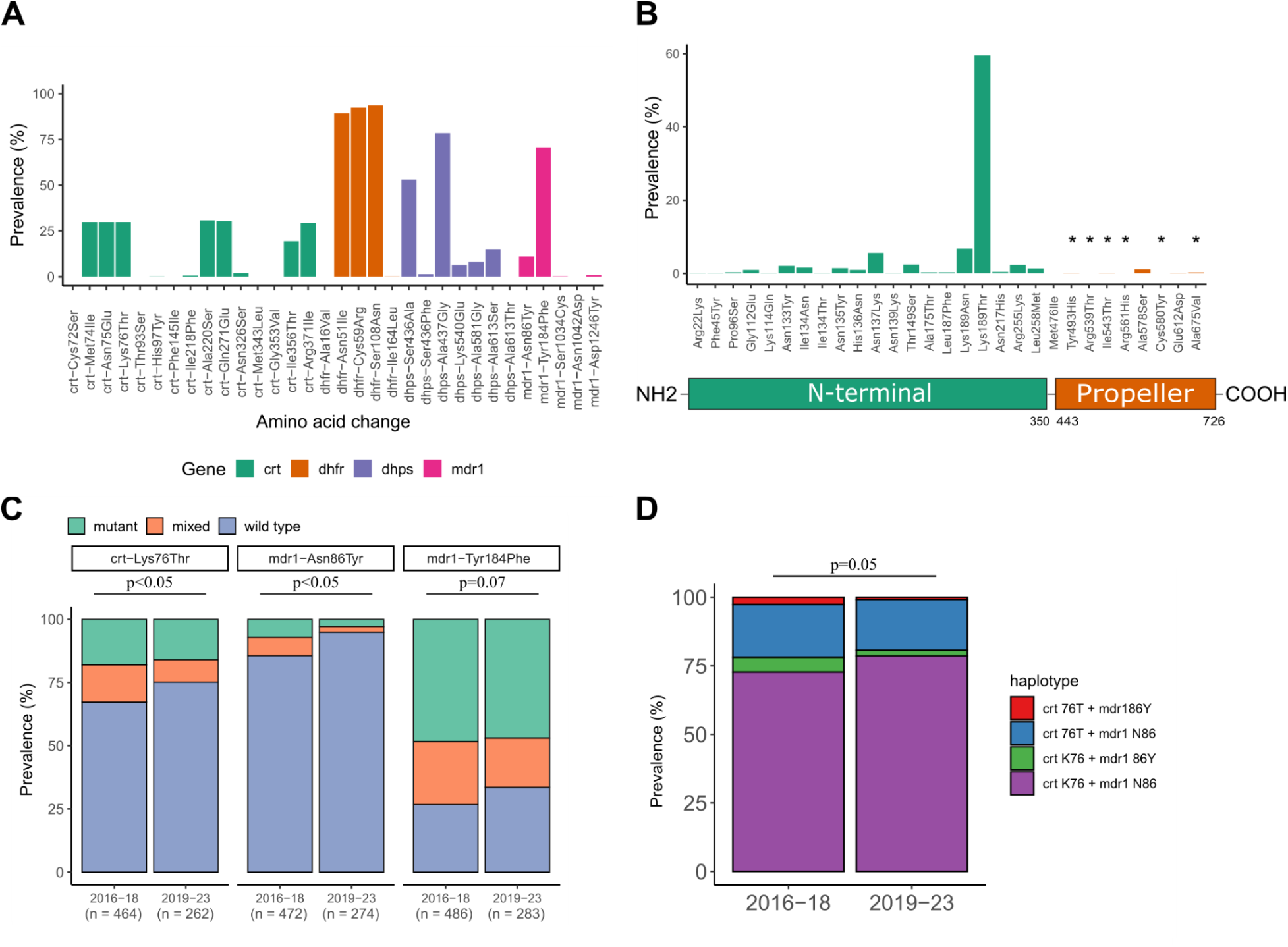
Prevalence of key mutations associated with resistance to different drugs. **A**) Prevalence of key mutations in *pfcrt*, *pfdhfr*, *pfdhps* and *pfmdr1* genes detected in isolates from this study. **B)** Prevalence of mutations in the *pfkelch13* gene. Green bar plots indicate the prevalence of mutations in the N-terminal-coding domain, while orange bar plots indicate the prevalence of mutations in the propeller domain. SNPs marked with an asterisk (*) are validated or candidate SNPs by WHO. **C)** Prevalence of the main key mutations in *pfcrt* and *pfmdr1* over time. **D)** Prevalence of *pfcrt* 76-*pfmdr1* 86 haplotypes over time. Color code for the 4 haplotypes: purple, wild-type K76-N86; green, single mutant K76-86**Y**; blue, single mutant 76**T**-N86; red, double mutant 76**T**-86**Y**. P-values were calculated by Fisher’s exact test.

In *pfmdr1,* the prevalence of the N86**Y**, Y184**F,** D1246**Y** mutations were 11%, 71%, and 1%, respectively (**Fig 3A)**, and the prevalence of the NYD, N**F**D and **YF**D haplotypes at codons 86-184-1246 were 38·8%, 54·8% and 5·6%, respectively (**Figure S4**). Other minor haplotypes were found in less than 1% of isolates.

The *pfcrt* K76**T** mutation was found in 30% of isolates (**Figure 3A**) and was absent in the Central African Republic (**Figure S5**). Regarding the classical eight *pfcrt* codons 74-75-76-220-271-326-356-371, three main haplotypes were detected (**Figure S4**; per country, see **Table S10** and **Figure S6**): the wild-type MNKAQNIR (77%) and the mutants **IETSE**N**TI** (also known as Cam783; 15%) and **IETSE**NI**I** (also known as GB4; 6%).

Four isolates carried a validated *pfkelch13* mutation in the propeller domain (**Figure 3B**): A675**V** (2/734, from Rwanda in 2019 and Uganda in 2020), Y493**H** (1/705, from Mali in 2016), and I543**T** (1/731, from Côte d’Ivoire in 2018). The two isolates carrying the A675**V** mutation showed a susceptibility to DHA (IC_50_ = 1·78 nM and IC_50_ = 1·67 nM) larger than the upper quartile. The A675**V** isolate from Rwanda also had elevated IC_50_ values for LMF (IC_50_ = 41·57 nM) and MFQ (IC_50_ = 49·12 nM). Two other mutations were detected in the propeller domain: A578**S** (8 isolates) that is known as a polymorphism and E612**D** (1 isolate). Most of the other non-synonymous *pfkelch13* mutations occurred in the N-terminal-coding domain (**Figure 3B, Figure S7**), K189**T** being very frequent (58·4%) and also known as polymorphism.

Finally, the prevalence of “background” mutations associated with ART-R in South East Asia ^27^ was low: *pffd* D193**Y** at 0·27%, *pfmdr2* T484**I** at 0%, *pfpib7* C1484**F** at 0·16%, and *pfpp* V1157**L** at 0% (**Table S11**).

We then analysed mutation prevalence over time (**Figure 3C, Figures S8** and **S9,** and **Table S12)**. Remarkably, when compared to samples collected in 2016-2018, a significant decrease in prevalence was observed in 2019-2023 samples for *pfcrt* K76**T** (33% in 2016-2018 versus 25% in 2019-2023; p<0·05) and *pfmdr1* N86**Y** (14% versus 5%; p<0·001), and there was a nearly significant decrease for *pfmdr1* Y184**F** (73% versus 66%; p=0·07) (**Figure 3C**). The prevalence of the wild-type haplotype at *pfcrt* 76 and *pfmdr1* 86 (K76-N86) was largely dominant across the whole study period and slightly increased from 73% in 2016-2018 to 79% in 2019-2023 (p=0·05, **Figure 3D, Figure S8B**). Analysis of SNP-SNP correlation in isolates from Côte d’Ivoire and Cameroon indicated that *pfmdr1* N86Y and *pfcrt* K76T were not in linkage disequilibrium with each other (**Figure S10**).

### Association between SNPs and drug susceptibility

Genotype-phenotype association analysis across 362 SNPs and susceptibilities to 6 drugs showed several significant associations (**Figure 4, Tables S12** for the full list of associations, and **S13** and **S14** after correction for multiple testing). Most strikingly, multiple well-known SNPs in *pfcrt* were strongly associated with susceptibilities to multiple drugs (CQ, MDAQ, DHA, LMF) and were borderline significant after multiple testing correction for MFQ and PPQ. Mutant *pfcrt* alleles were associated with decreased susceptibility to CQ (p = 10^−38^−10^−50^ for 7 amino acids) and MDAQ (p = 10^−6^−10^−8^ for 7 amino acids) whereas wild-type *pfcrt* alleles were associated with decreased susceptibility to DHA (p = 10^−6^ for 6 amino acids) and LMF (p = 10^−5^ for 3 amino acids). There was also a strong association between the well-characterised mutation N86**Y** in *pfmdr1* and susceptibility to MFQ (p = 10^−6^), LMF (p = 10^−5^), and DHA (p=0·0003, although non-significant after correction). Finally, the *pfubp1* N1704**K** and K1705**N** mutations were associated with decreased susceptibility to CQ (p = 2·10^−5^ and 0·0001 respectively).

**Figure 4.**
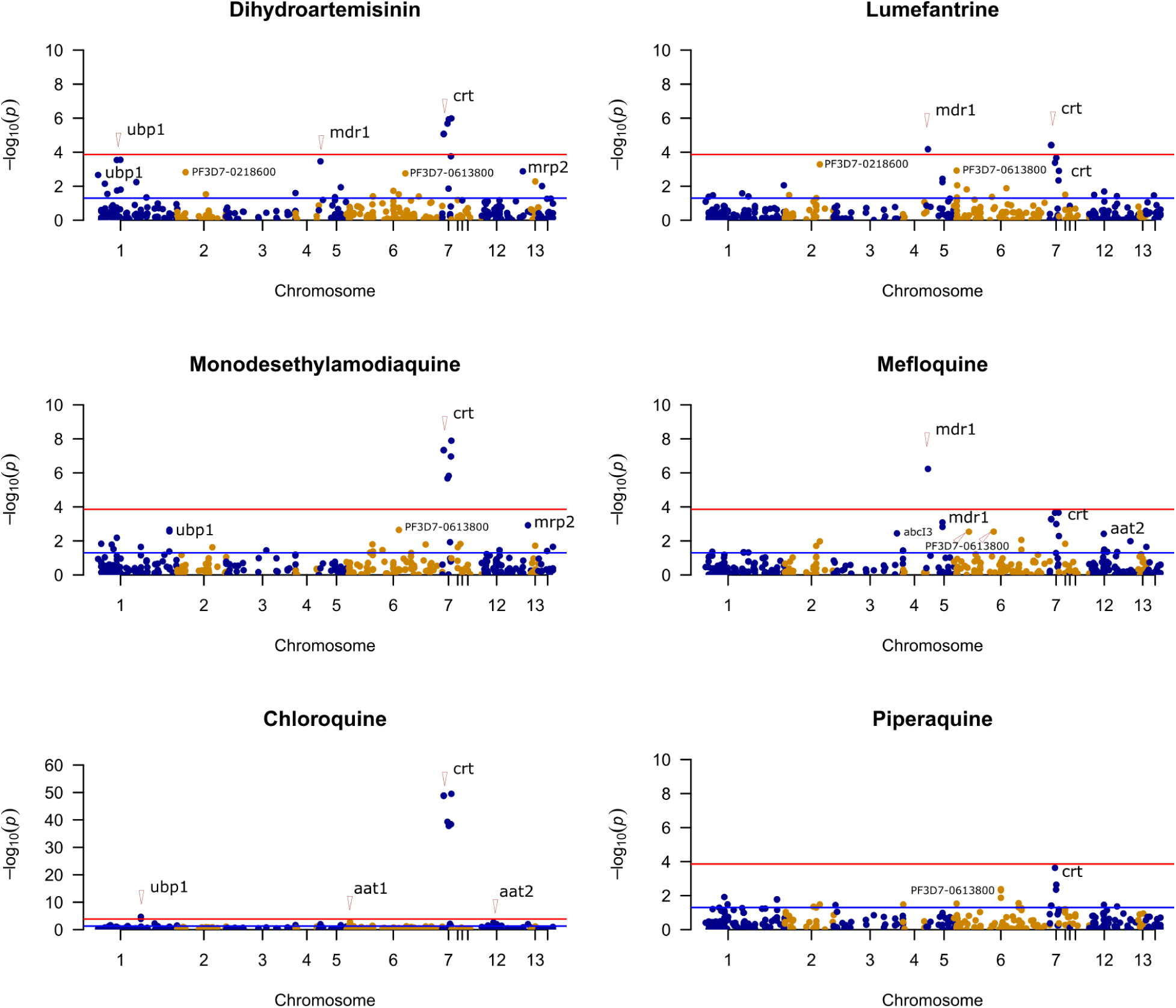
Manhattan plot showing the significance of SNPs associated with six antimalarial drugs. Each dot represents 1 of 362 SNPs with MAF>0·01 colored by chromosome. The x-axis represents the chromosomal location of the SNPs, and the y-axis represents the –log10 of the P-value obtained from the linear regression model analysis. The blue line represents the nominal P-value (P ≤ 0·05), and the red line represents the P-value after Bonferroni correction (P ≤ 1×10^−4^). The full list of SNPs associated with IC50 is shown in **Table S13**.

To control for the major *pfcrt* effect, we added *pfcrt* K76T as a covariate (**Table S13**). All the associations involving another SNP in *pfcrt* (except M74I for CQ) and *pfubp1* N1704K and K1705N were lost, suggesting they were dependent on the *pfcrt* K76T genotype. The associations involving *pfmdr1* (with LMF and MFQ) were maintained and no new associations were detected. A model with *pfmdr1* N86Y as a covariate abolished only the association of *pfubp1* K1705N for CQ (but not the one of N1704K) (**Table S13**). These data indicate that, among the 362 tested SNPs, *pfcrt* and *pfmdr1* SNPs were the major drivers of susceptibility to most tested drugs and could interact in a likely additive manner in the case of LMF.

### Effect of *pfcrt* and *pfmdr1* haplotypes on IC_50_s

As compared to the wild-type *pfcrt* (MNKAQNIR), mutant haplotypes **IETSE**N**TI** and **IETSE**NI**I** (that differ at I356**T** only) were significantly associated with decreased susceptibility to CQ and MDAQ (Kruskal-Wallis test, p-value range: <0·0001-0·0003; **Figure S11**) and improved susceptibility to DHA, LMF and PPQ (Kruskal-Wallis test, p-value range: <0·0001-0·05). **IETSE**N**TI**, but not **IETSE**NI**I,** was associated with improved susceptibility to MFQ (Kruskal-Wallis test, p = 0·0001), suggesting that I356**T** has some role in MFQ transport activity.

Isolates carrying the *pfmdr1* NYD wild-type haplotype had decreased susceptibility to DHA, LMF, MFQ and improved susceptibility to PPQ as compared to the mutant **YF**D (Kruskal-Wallis test, p-value range: <0·001 – 0·01; **Figure S11**).

Finally, the analysis of *pfcrt* K76T – *pfmdr1* N86Y haplotypes identified three main drug patterns (**Figure 5**). The susceptibility to LMF, MFQ, and also marginally to DHA, improved with the number of *pfcrt-pfmdr1* mutations suggesting these mutations work in additive ways. The most decreased susceptibility to LMF was associated with the wild-type haplotype (*pfcrt* K76-*pfmdr1* N86). The susceptibility to CQ and MDAQ decreased mostly with mutant *pfcrt*, and the addition of mutant *pfmdr1* had a lesser effect. The susceptibility to PPQ was more difficult to interpret with little haplotypic effects.

**Figure 5.**
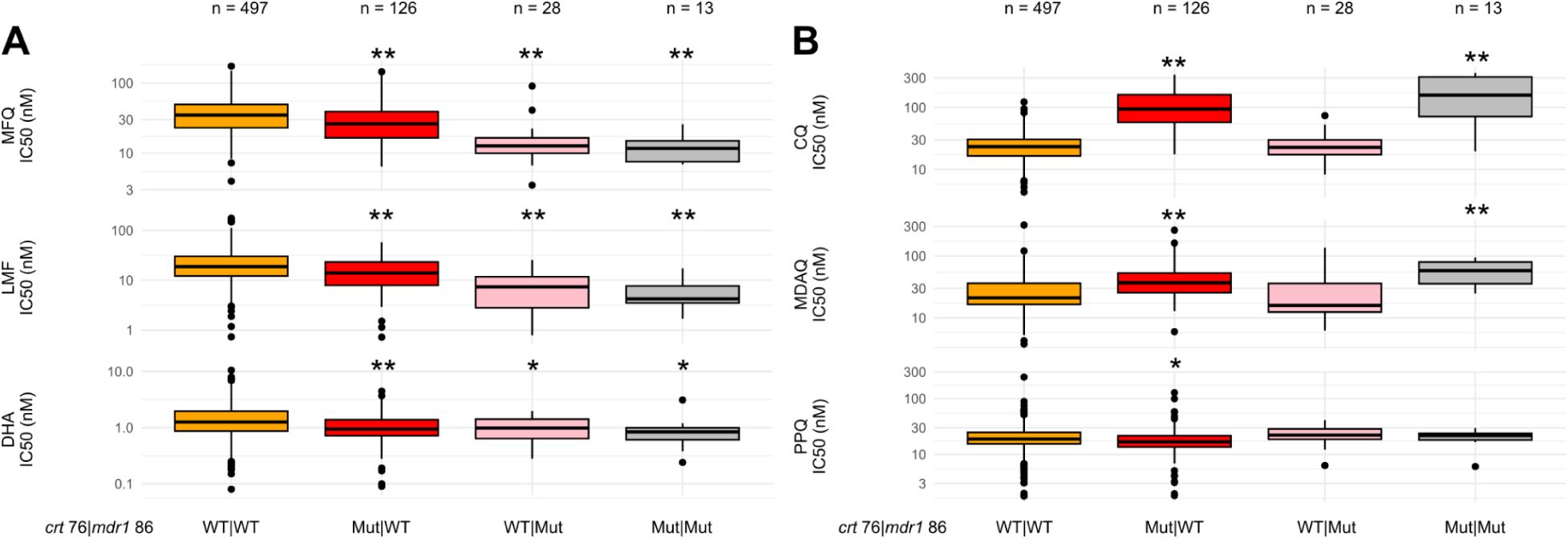
Effect of *pfcrt* 76-*pfmdr1* 86 haplotypes on IC_50_ for six drugs. **A**) IC_50_ for mefloquine (MFQ), lumefantrine (LMF) and dihydroartemisinin (DHA) disaggregated by *pfcrt-pfmdr1* haplotypes. B) IC_50_ for chloroquine (CQ), monodesethylamodiaquine (MDAQ) and piperaquine (PPQ) disaggregated by *pfcrt-pfmdr1* haplotypes. Haplotypes were built with *pfcrt* K76T and *pfmdr1* N86Y using the major allele per position. n: number of isolates carrying the haplotype. Differences in the IC_50_ of wild-type (WT|WT, crt-K76|mdr1-N86) versus single mutants (Mut|WT, crt-76T|mdr1-N86), (WT|Mut, crt-K76|mdr1-86Y), and double mutant (Mut|Mut, crt-76T|mdr1-86Y) were calculated with a Pairwise Wilcoxon tests with Benjamini-Hochberg correction. *: p<0·05; **: p<0·001.

## Discussion

Whereas there are recent reports of emerging ART-R and decreased susceptibility to LMF in *P. falciparum* parasites from East Africa, ^28^ few recent studies have investigated the resistance status of contemporary West and Central African isolates at a large scale. Here, we report the temporal trends of *ex vivo* susceptibilities to six antimalarials using standard growth inhibition assays and their association with mutations in fourteen drug resistance genes in 805 *P. falciparum* samples collected between 2016 and 2023 in returned travelers to West and Central African countries. We found an overall excellent susceptibility of isolates to the six evaluated antimalarials, but the one to ACT partner drugs LMF and MDAQ decreased in recent years. Concomitantly, the prevalence of wild-type *pfcrt* K76 and *pfmdr1* N86 alleles increased in recent years, consistent with a large selective pressure exerted by the use of LMF and the discontinuation of CQ in these regions. Finally, altered susceptibility to most drugs was strongly associated with *pfcrt* and/or *pfmdr1* genotypes.

Regarding molecular ART-R, only four isolates among 805 carried a WHO-validated *pfkelch13* mutation. A675**V** was detected in two travelers returning from East Africa (Rwanda and Uganda), consistent with the spread of this mutation in this region.^10,29^ The returned traveler from Uganda showed delayed clearance under artesunate therapy,^30^ whereas the A675**V** isolate from Rwanda, had elevated IC_50_ values for LMF (41.57 nM) and MFQ (49.12 nM), consistent with clinical and *ex vivo* data reported by others for Ugandan A675**V** isolates.^11,14,15^ Two other validated mutations, Y493**H** and I543**T**, were identified in one isolate from Mali in 2016 and one from Côte d’Ivoire in 2018, respectively. These isolates did not have unusual IC_50_s against any of the drugs tested. As these two mutations were not detected in more recent samples, these seem to be sporadic cases. Altogether, *pfkelch13* ART-R mutations were detected but uncommon in Central and West African isolates during the study period (2016-2023). Contrary to the current situation in East Africa, *pfkelch13*-mediated ART-R is seemingly not emerging during 2016-2023 in West and Central African regions. Continuous monitoring of *pfkelch13* mutations should however be reinforced as they could either emerge locally or be introduced in these regions by intracontinental migration from East Africa.

We found a significant reduction in susceptibility of *P. falciparum* to LMF and MDAQ for samples collected in recent years. LMF and MDAQ are the two main partner drugs used in West and Central Africa during the last decade. The use of ACTs may have selected parasites whose response to the partner’s drug is altered in these regions. A similar trend was previously reported for LMF in northern Uganda,^14^ which possibly predated the emergence of ART-R in this region. ^10^ The clinical effects of the decreased susceptibilities to LMF and MDAQ observed in our study are unknown, but the spread of ART-R may be facilitated in such phenotypic backgrounds.

Remarkably, wild-type *pfcrt* and *pfmdr1* genes were individually associated with decreased susceptibility to LMF, consistent with previous *in vitro* and clinical findings.^31,32,33^ Haplotypic association analysis further suggests that the two genes have cumulative effects, with the wild-type haplotype (*pfcrt* K76**-***pfmdr1* N86) exhibiting the largest decrease in susceptibility to LMF. The increasing prevalence in recent years of parasites carrying the wild-type allele at either gene or at both could, therefore, explain the LMF data. In contrast the double mutant haplotype (*pfcrt* 76T**-***pfmdr1* 86Y) exhibited the largest decrease in susceptibility to MDAQ. Taken together, these findings support the rationale for implementing regimens with drug combinations that exert opposing selective pressures.^34^ Of note, susceptibility to MDAQ decreased in recent years, whereas the prevalence of the *pfcrt* K76**T** mutation decreased slightly. This is surprising as this mutation is strongly associated with decreased susceptibility to MDAQ in our dataset and other studies. We have no solid explanation for this result, but one possibility could be that other mediators of altered susceptibility to MDAQ exist and that our targeted MIP sequencing and association studies did not capture them.

Intriguingly, two novel *pfubp1* mutations, N1704**K** and K1705**N,** were associated with reduced susceptibility to CQ in our data set. The effect of these *pfubp1* mutations seems to depend on the *pfcrt* K76**T** genotype. However, as these two mutations have low prevalence (10.0 and 7.9%, respectively) and likely a modest effect, we have low statistical power to conduct a haplotypic analysis. Several studies reported an association of different *upb1* mutations with altered survival or susceptibility to DHA in mouse malaria or *P. falciparum*,^35,14^ and recently to MFQ, LMF and PPQ in the *Plasmodium yoelii* mouse malaria model.^36^ However, as the two new mutations are located in an N/K repeat protein region that is not part of the reported active site(s) of the enzyme, gene editing experiments are needed to formally address their role in modulating drug response to CQ.^37^

The high prevalence of pyrimethamine resistance mutations in the study period indicates the high level of SP pressure in West and Central Africa. Nevertheless, the low prevalence of *pfdhps* K540E (<10%) and of the SP-resistant quintuple *dhfr-dhps* mutant (3%) is reassuring that SP is still effective for preventive therapies. Indeed, the WHO recommends for countries to withdraw SP for IPTp use when the prevalence of *Pfdhps* K540E>95% and A581G>10% and for IPTi use when the prevalence of *Pfdhps* K540E >50%.^38^

This study was not conducted without limitations. First, we have not evaluated *ex vivo* resistance to artemisinin derivatives with the dedicated ring survival assay, the standard measure of in vitro susceptibility to artemisinins. We, however, report very rare ART-R mutations in *pfkelch13,* and at this time, these are the major determinants of ART-R in patients. Second, we have not evaluated copy number variations in *pfmdr1* and *pfplasmepsins 1-2* that are associated with altered susceptibility to MFQ and PPQ, respectively. Third, our geographical coverage of West and Central Africa is biased, with two countries contributing to almost 50% of the total isolate sampling (169 isolates from Cameroon in Central Africa and 217 isolates from Côte d’Ivoire in West Africa). Therefore, trends at the country level could not be robustly drawn. Fourth, our analyses focused on imported malaria cases to France and infections in travelers may differ in several aspects from those in people living in endemic areas. Finally, we used a targeted sequencing approach to genotype isolates and focused on validated and some candidate drug resistance genes. Detecting emerging resistance mutations in novel genes would need a larger genomic coverage.

In conclusion, our study provides much-needed information on molecular and phenotypic resistance in contemporary isolates from West and Central Africa. It shows how current and past control efforts have been affecting malaria parasite populations in those regions and calls for intensifying the monitoring to inform as quickly as possible on future changes regarding antimalarial drug susceptibilities and resistance.

## Author contributions

JR, AAF, SC, VS, RCo, JC, JAB, and SH designed the study. SC, VS, RZ, AB, LC, and LH did the *ex-vivo* IC_50_ assays, and archived data. LM, MT, BP, JC and SH provided administrative and logistical support. JR, AAF, JC, RCr, and JAB analysed and did the genotyping. JR and AAF verified and analysed the data and did the statistical analysis. All authors had full access to all the data. All authors contributed to the writing of the manuscript.

## Acknowledgements

We thank the participants who anonymously agreed to participate in a malaria drug resistance surveillance study in France, the staff at the CNR for their support in collecting isolates and executing the drug resistance test, Justine Bailly at MERIT unit, and Alec Leonetti’s MIP sequencing support at Brown University. The members of the French National Reference Centre for Imported Malaria Study Group are as follows: Dr A. Aboubacar (CHU Strasbourg), Dr P. Agnamey (CHU Amiens), Dr A. Ahmed (CHU Strasbourg), Dr D. Ajzenberg (CHU Limoges), Dr A. Angoulvant (CHU Bicetre), Dr F. Ariey (CHU Cochin, Paris), Dr B. Aubry (CH Le Mans), Pr F. Banisadr (CHU Reims), Dr S. Belaz (CHU Pontchaillou Rennes), Dr G. Belkadi (CHU Saint Antoine, Paris), Dr A. Bellanger (CHU Jean Minjoz,-Besancon), Dr D. Bemba (CHU Jean Verdier, Bondy), Dr F. Benaoudia (CH Troyes), Dr J. Benjamin Murat (CH Roanne), Dr M. Bloch (CHU Louis Mourier, Colombes), Dr F. Botterel (CHU Henri Mondor, Créteil), Dr V. Bouden (CH Saint-Nazaire), Dr M. Bougnoux (CHU Necker Enfants Malades, Paris), Dr C. Brump (CHU Lariboisière, Paris), Dr S. Brun (CHU Avicenne, Bobigny), Dr J. Brunet (CHU Strasbourg), Dr B. Buret (CH Niort), Dr P. Caraux-Paz (CH Villeneuve St Georges), Dr N. Celine (CHU Clermont Ferrand), Dr A. Cherif Touil (CH Niort), Dr S. Clauser (CHU Ambroise Paré, Boulogne Billancourt), Dr E. Collin (CH R Ballanger, Aulnay sous-bois), Dr N. Dahane (CHU Cochin, Paris), Dr C. Damiani (CHU Amiens), Dr E. Dannaoui (CHU HEGP, Paris), Dr C. Dard (CHU Grenoble), Pr M. Darde (CHU Limoges), Dr L. De Gentile (CHU Angers), Dr A. Debourgogne (CHU Nancy), Dr T. Delacour (CH Creil), Dr J. Delarbre (CH Mulhouse), Dr A. Delaval (CH R Ballanger, Aulnay sous-bois), Dr V. Delcey (CHU Lariboisière, Paris), Dr A. Deleplancque (CHU Lille), Dr G. Desoubeaux (CHU Bretonneau, Tours), Dr G. Desoubeaux (CHU Tours), Dr N. Desuremain (CHU Armand Trousseau, Paris), Dr G. Dewulf (CH Valenciennes),Dr R. Durand (CHU Avicenne, Bobigny), Dr M. Durieux (CHU Limoges), Dr E. Dutoit (CHU Lille), Dr M. Edith (CH Valenciennes), Dr O. Eloy (CH André Mignot, Le Chesnay), Dr M. Evrard Sylvie (CH Villeneuve St Georges), Dr J. Faucher (CHU Limoges), Pr L. Favennec (CHU Ch,Pr A. Faye (CHU Robert Debre, Paris), Dr O. Fenneteau (CHU Robert Debre, Paris), Dr C. Ficko (Hia Begin, Saint-Mandé), Dr E. Frealle (CHU Lille), Dr R. Gabrielle (CH Tourcoing), Dr G. Gargala (CHU Rouen), Dr C. Garnaud (CHU Grenoble), Dr N. Godineau (CH Delafontaine, Saint-Denis), Dr J. Gorlicki (CHU Lariboisière, Paris), Dr A. Gravet (CH Mulhouse), Dr N. Guennouni (CHU Bicetre, Le Kremlin Bicêtre), Dr S. Hamane (CHU Saint-Louis, Paris), Dr T. Hanslik (CHU Ambroise Paré, Boulogne Billancourt), Dr A. Huguenin (CHU Reims), Dr J. Hurst (CH Jacques Monod, Le Havre), N. Imbert (APHP, Hôpital Bichat-Claude-Bernard), Dr F. Jeddi (CHU Nantes), Dr L. Jordan (CHU Lille), Dr L. Landraud (CHU Louis Mourier, Colombes), Dr H. Lapillonne (CHU Armand Trousseau, Paris), Dr S. Larréché (Hia Begin, Saint-Mandé), Dr R. Lavergne (CHU Nantes), Dr Y. Le Govic (CHU Angers), Dr G. Le Moal (CHU Poitiers), Dr J. Lemoine (CHU Angers), Dr C. Leprince (CH R Ballanger, Aulnay sous-bois), Dr E. Lesteven (CHU Lariboisière, Paris), Dr A. Li (CH Creil), Dr C. Lohmann (CH Mulhouse), Dr B. Louise (CHU Dijon), Dr M. Machouart (CHU Nancy), Dr C. Malassigne (CH Le Havre), C. Maréchal (APHP, Hôpital Bichat-Claude-Bernard), Dr A. Marteau (CHU Avicenne, Bobigny), Dr D. Maubon (CHU Grenoble), Dr E. Mazars (CH Valenciennes), Pr B. Megarbane (CHU Lariboisière, Paris), Dr A. Mendes-Moreira (CH La Rochelle), Dr S. Mermond (GH La Rochelle), C. Moissant, (APHP, Hôpital Bichat-Claude-Bernard), Dr A. Moreno (CHU Saint Antoine, Paris), Dr P. Mornand (CHU Armand Trousseau, Paris), Dr J. Murat (CHU Limoges), Dr R. Nabias (CH Poissy/Saint-Germain-En-Laye), Dr J. Naudin (CHU Robert Debre, Paris), Dr G. Nevez (CHU Brest), Pr G. Nevez (CHU La Cavale Blanche, Brest), Dr C. Nourrisson (CHU Clermont-Ferrand), Dr M. Oussama (CHU Pitié Salpetrière, Paris), Dr N. Dr P. Patoz (CH Tourcoing), Pean-De-Ponfilly (CHU Lariboisière, Paris), Dr J. Peltier (CH Poissy/Saint-Germain-En-Laye), Dr P. Penn (CH Le Mans), Dr A. Perignon (CHU Pitié Salpetrière, Paris), Dr E. Perraud-Cateau (CHU Poitiers), Dr A. Pfaff (CHU Strasbourg), Dr M. Pihet (CHU Angers), Dr P. Poirier (CHU Clermont Ferrand), Dr J. Pilo (Hia Begin, Saint-Mandé), Dr I. Poilane (CHU Jean Verdier, Bondy), Dr D. Poisson (CH Orléans), Dr D. Pons (CHU Clermont Ferrand), Dr L. Pull (CHU Robert Debre, Paris), Dr D. Quinio (CHU Brest), Dr D. Raffenot (CH Chambery), Dr M. Revest (CHU Pontchaillou, Rennes), Dr G. Robert (CHU Grenoble), Dr O. Rogeaux (CH Chambery), Dr M. Sasso (CHU Caremeau Nimes), Dr F. Schmitt (CH Mulhouse), Dr B. Sendid (CHU Lille), Dr Y. Senghor (Groupe Hospitalier Saint Joseph, Paris), Dr M. Silva (CH Jacques Monod, Le Havre), Dr J. Siriez (CHU Robert Debré, Paris), Dr E. Sitterlé (CHU Necker Enfants Malades, Paris), Dr F. Sorge (CHU Necker Enfants Malades, Paris), Dr I. Tantaoui (CHU Pitie-Salpetriere, Paris), G. Thaboulet (APHP, Hôpital Bichat-Claude-Bernard), Dr D. Toubas (CHU Reims), Dr C. Tournus (CH Saint-Denis), L. Wallus, (APHP, Hôpital Bichat-Claude-Bernard), Dr H. Yera (CHU Cochin, Paris).

## Data availability

All sequencing data are available under accession no. SAMN42141774 to SAMN42142576 at the Sequence Read Archive (SRA), and the associated BioProject ID is PRJNA1129163. De-identified datasets generated during the current study and used to make all figures are available as supplementary files or tables. The data and R scripts developed in this study were deposited in the GitHub repository (https://github.com/jrosados/CNR_IC50_MIPs). Additional software packages and tools that are useful when working with MIP data are available at https://github.com/bailey-lab/MIPTools and https://github.com/Mrc-ide/mipanalyzer.

