## Supplementary appendix for "*Ex vivo* susceptibility to antimalarial drugs and polymorphisms in drug resistance genes of African *Plasmodium falciparum*, 2016-2023: a genotype-phenotype association study": Supplementary_material_EditME_updated_120724.pdf

**Supplementary appendix 1**

### *Supplemental to: Assessment of ex-vivo antimalarial drug efficacy in African Plasmodium falciparum parasite isolates, 2016-2023: a genotype-phenotype association study*

Jason Rosado[1]*¥, Abebe A. Fola[2,3]¥, Sandrine Cojean[4,5,6]¥, Véronique Sarrasin[4]¥, Romain Coppée[7], Rizwana Zaffaroulah[4], Azza Bouzayene[4], Liliane Cicéron[4], Céline Maréchal[4], Geoffrey Thaboulet[4], Camille Moissant[4], Léa Wallus[4], Ludivine Houzé[4], Nicolas Imbert[4], Rebecca Crudale[2,3], Lise Musset[8], Marc Thellier[9], Bruno Pradines[10,11,12,13], Jérôme Clain[1]*¶, Jeffrey A. Bailey[2,3]¶, Sandrine Houzé[1,4]¶, Investigation Study Group[Ψ]

Authors and affiliations

[1] Université Paris Cité, IRD, MERIT, F-75006 Paris, France
[2] Department of Pathology and Laboratory Medicine, Brown University, RI, USA, 02906
[3] Center for Computational Molecular Biology, Brown University, RI, USA, 02906
[4] Centre National de Référence du Paludisme, Assistance Publique-Hôpitaux de Paris, Hôpital Bichat-Claude-Bernard, Paris, France
[5] Université Paris-Saclay, Faculté de Pharmacie, Orsay, France
[6] UMR BIPAR, Anses, Laboratoire de Santé Animale, INRAE, Ecole Nationale Vétérinaire d'Alfort, Maisons-Alfort, France
[7] Université de Rouen Normandie, Laboratoire de Parasitologie-Mycologie, UR 7510 ESCAPE, Rouen, France
[8] Laboratoire de Parasitologie, World Health Organization Collaborating Center for Surveillance of Antimalarial Drug Resistance, Centre Nationale de Référence du Paludisme, Institut Pasteur de la Guyane, Cayenne, French Guiana.
[9] Centre National de Référence du Paludisme, Sorbonne Université, Assistance Publique des Hôpitaux de Paris, Laboratoire de Mycologie et Parasitologie, Hôpital de la Pitié-Salpêtrière, Paris, France.
[10] Unité Parasitologie et Entomologie, Département des Maladies Infectieuses, Institut de Recherche Biomédicale des Armées, Marseille, France
[11] Aix Marseille Université, SSA, AP-HM, RITMES, Marseille, France
[12] Centre National de Référence du Paludisme, Marseille, France
[13] IHU-Méditerranée Infection, Marseille, France

### Supplementary appendix 1


**Supplementary Methods (pages: 2-3)**

*Ex-vivo* assays

MIP data analysis and estimating drug resistance prevalence.

**Supplementary Tables (pages: 4-23)**

**Table S1.** List of drug resistance markers across *Plasmodium falciparum* genome included in drug resistance MIP panel.

**Table S2.** List of countries visited by travelers returning to France over 2016-2023.

**Table S3.** Frequency of isolates from West and Central Africa region in 2016-18 and 2019-23.

**Table S4.** Drug susceptibility of control strain 3D7.

**Table S5.** Drug susceptibility in 2016-18 vs 2019-23.

**Table S6.** Drug susceptibility in 2016-18 versus 2019-23 stratified by African region.

**Table S7.** Drug susceptibility and temporal trends over time.

**Table S8.** Frequency of resistant isolates defined by cut-off in 2016-18 and 2019-23.

**Table S9.** Prevalence (%) of key mutations in the top ten countries visited by participants of this study from 2016-2023.

**Table S10.** Frequency distribution of *pfcrt* haplotypes in the ten top countries visited by participants of this study.

**Table S11.** Prevalence of ART-R background mutations.

**Table S12.** Prevalence (%) of key mutations by year in all isolates.

**Table S13.** Full list of mutations associated with $IC_{50}$ identified by targeted genome association tests.

**Table S14.** Genomic loci associated with $IC_{50}$ identified by targeted genome association tests.

**Table S15.** Distribution of $IC_{50}$ according to associated SNPs.

**Supplementary Figures (pages: 24-34)**

**Figure S1.** Scatter plot of $IC_{50}$ for Dihydroartemisinin and partner drugs indicating $IC_{50}$ cut-offs.

**Figure S2.** Correlation between pairs of $IC_{50}$ values.

**Figure S3.** MIP sequencing coverage- complexity of infections and parasitemia of isolates.

**Figure S4.** Pie charts of *pfdhfr*- *pfdhps*- *pfmdr1* and *pfcrt* haplotypes.

**Figure S5.** Prevalence of key mutations in the ten most visited countries by participants.

**Figure S6.** Frequency distribution of the *pfcrt* haplotypes in the ten most visited countries by participants.

**Figure S7.** Prevalence of mutations in *pfkelch13* gene in the ten most visited countries by participants.

**Figure S8.** Temporal change of key resistance mutations and haplotypes by year.

**Figure S9.** Prevalence of key resistance mutations by year and by country.

**Figure S10.** Linkage disequilibrium of SNPs associated with drug resistance in isolates from travelers to Cameroon and Ivory Coast.

**Figure S11.** Effect of *pfcrt* and *pfmdr1* haplotypes in $IC_{50}$ for six drugs.


### Supplementary appendix 2

Supplementary Appendix 2 (worksheets 1-3; Excel file: Appendix_2.xlxs)

### Supplementary methods

The geographic distribution of traveler isolates was plotted using the R packages rnaturalearth (v.1.0.1) and rnaturalearthdata (v.1.0.0) in R version 4.3.1.

***Ex-vivo* assays.** Chloroquine (CQ)- dihydroartemisinin (DHA)- mono-desethyl-amodiaquine (MDAQ)- MFQ- LMF- and PPQ were purchased from Alsachim (Illkirch Graffenstaden- France). Briefly- MDAQ- MFQ- and DHA were first dissolved in methanol and then diluted in water to final concentrations ranging from 1.9 to 2-000 nM for MDAQ- 1.5 to 400 nM for MFQ- and 0.1 to 100 nM for DHA. CQ was diluted in water to final concentrations ranging from 6.38 to 3-300 nM. LMF was resuspended and diluted in ethanol to final concentrations ranging from 0.6 to 310 nM. PPQ was diluted in 0.5% lactic acid and then diluted in water to final concentrations ranging from 2 to 1-000 nM. The susceptibility of the isolates to the antimalarial drugs was assessed without culture adaptation. A total of 100 µL of parasitized erythrocytes (final parasitaemia of 0.5% and a final hematocrit of 1.5%) were aliquoted into 96-well plates preloaded with a concentration gradient of antimalarial drugs. The plates were incubated for 48 hours in a controlled atmosphere of 85% $N_2$- 10% $O_2$- 5% $CO_2$ at 37°C. Drug susceptibility was determined using the standard 42-hour [$^3$H]hypoxanthine uptake inhibition assay[1]. Each batch of plates was validated using the CQ-sensitive reference strain 3D7 (Africa) and the CQ-resistant reference strain W2 (Indochina) in independent experiments under the same conditions described below. The 50% inhibitory concentration ($IC_{50}$) was estimated using the non-linear regression ICEstimator[2]. The cut-off values for reduced *in vitro* susceptibility or resistance were as follows: 10 nM (DHA)- 150 nM (LMF)- 30 nM (MFQ)- 100 nM (CQ)- 80 nM (MDAQ)- and 135 nM (PPQ)[3–5].

**MIP data analysis and estimating drug resistance prevalence.** Sequencing data processing and variant calling were performed using MIPtools (v0.19.12.13; https://github.com/bailey-lab/MIPTools)- a suite of computational tools designed to handle sequencing data from MIPs. Using MIPWrangler- raw reads from each captured MIP- identifiable by unique molecular identifiers (UMIs)- were used to reconstruct sequences- and variant calling was performed on these samples using freebayes[6]. Biallelic- variant SNP positions were retained for analysis. Variants were annotated using the 3D7 v3 reference genome. To reduce false positives due to PCR and sequencing errors- the alternative allele (SNP) must have been supported by more than one UMI within a sample with at least 5x coverage- and the allele must have been represented by at least 10 UMIs in the population.

We estimated the prevalence of validated mutations as reported by the WHO. In addition- the prevalence of reported “background mutations”[7] associated with ART-R such as *ferredoxin* (*pffd*) D193Y- *multidrug resistance 2 transporter* (*pfmdr2*) T484I- *putative phosphoinositide binding protein* (*pfpib7*) C1484F- *protein phosphatase* (*pfpp*) V1157L- and *pfcrt* N326S and I356T was also assessed. The prevalence for each drug resistance marker was calculated as $p = x / n * 100$- where p is the prevalence- x is the number of infections with mutant alleles- and n is the number of successfully genotyped infections- as previously described[8]. Mixed genotypes- i.e. a sample with a reference allele and an alternative allele in a given sample- were considered mutant regardless of the within-sample mutant allele frequency.

Haplotypes were reconstructed for all samples using the major allele in a given codon- i.e. the major allele was the allele with a within-sample allele frequency >75%; alleles with a frequency between 25% and 75% were considered unresolved and discarded from the analysis. Only samples with complete SNP data for the tested haplotypes were included in the analysis.

**Phenotype–genotype association analysis.** We identified 3,439 exonic mutation types, including 1041 indels and 2,398 SNPs across the 43 exons from 14 genes reported to be associated with drug resistance. Isolates with >30% missing genotype data were discarded. We then retained only biallelic non-synonymous SNPs with minor allele frequency (MAF) >0.01 for phenotype-genotype association analysis. After filtering, we retained 362 biallelic exonic SNPs with minor allele frequency (MAF) >0.01 across 805 isolates.

Phenotype-genotype association was performed with the SNPassoc R package (v.2.1.0)[9] using the built-in WGassociation function under a dominant model. For each drug, a linear regression model was fitted with continuous $IC_{50}$ values as the outcome and individual SNP genotypes as independent variables. The model was adjusted for the complexity of infection (COI), year, and the visited country of imported cases as covariates. We applied the Bonferroni correction to define a significance threshold of $P \leq 1\times10^{-4}$ for all analyses. Association analysis results were visualized with a Manhattan plot using the qqman R package (v.0.1.9). COI was estimated using the R package RealMcCoil (v.1.3.1)[10].

As a secondary analysis, conditional regression using *pfcrt* SNPs was performed to remove any effect of *pfcrt* loci and to detect novel variants associated with drug susceptibility.

To investigate linkage disequilibrium between drug resistance markers at *pfdhfr*, *pfmdr1*, *pfcrt*, and *pfdhps* loci, we used biallelic SNPs with MAF > 0.01 and within-sample majority alleles. Linkage disequilibrium analysis was plotted using LDheatmap (v.1.0.5)[11].

**Table S1. List of drug resistance markers across *Plasmodium falciparum* genome included in drug resistance MIP panel.**

| Chromosome | Genomic position | Reference Allele | Alternative allele | Gene ID | Gene Name | Aminoacid Change Position | Aminoacid Change | Mutation Names |
|---|---|---|---|---|---|---|---|---|
| chr1 | 267306 | G | T | PF3D7_0106300 | atp6 | A623 | Ala623Glu | atp6-Ala623Glu |
| chr1 | 268386 | AA | TC | PF3D7_0106300 | atp6 | L263 | Leu263Glu | atp6-Leu263Glu |
| chr1 | 267883 | C | T | PF3D7_0106300 | atp6 | E431 | Glu431Lys | atp6-Glu431Lys |
| chr1 | 266868 | C | T | PF3D7_0106300 | atp6 | S769 | Ser769Asn | atp6-Ser769Asn |
| chr4 | 748239 | A | T | PF3D7_0417200 | dhfr-ts | N51 | Asn51Ile | dhfr-ts-Asn51Ile |
| chr4 | 748577 | A | T | PF3D7_0417200 | dhfr-ts | I164 | Ile164Leu | dhfr-ts-Ile164Leu |
| chr4 | 748410 | G | A | PF3D7_0417200 | dhfr-ts | S108 | Ser108Asn | dhfr-ts-Ser108Asn |
| chr4 | 748262 | T | C | PF3D7_0417200 | dhfr-ts | C59 | Cys59Arg | dhfr-ts-Cys59Arg |
| chr4 | 748410 | G | C | PF3D7_0417200 | dhfr-ts | S108 | Ser108Thr | dhfr-ts-Ser108Thr |
| chr4 | 748134 | C | T | PF3D7_0417200 | dhfr-ts | A16 | Ala16Val | dhfr-ts-Ala16Val |
| chr4 | 881071 | A | G | PF3D7_0419900 | PI4K | S915 | Ser915Gly | PI4K-Ser915Gly |
| chr5 | 960989 | A | T | PF3D7_0523000 | mdr1 | S1034 | Ser1034Cys | mdr1-Ser1034Cys |
| chr5 | 961013 | A | G | PF3D7_0523000 | mdr1 | N1042 | Asn1042Asp | mdr1-Asn1042Asp |
| chr5 | 958440 | A | T | PF3D7_0523000 | mdr1 | Y184 | Tyr184Phe | mdr1-Tyr184Phe |
| chr5 | 961625 | G | T | PF3D7_0523000 | mdr1 | D1246 | Asp1246Tyr | mdr1-Asp1246Tyr |
| chr5 | 958145 | A | T | PF3D7_0523000 | mdr1 | N86 | Asn86Tyr | mdr1-Asn86Tyr |
| chr6 | 1066989 | A | G | PF3D7_0626400 | Sec14 | N615 | Asn615Asp | Sec14-Asn615Asp |
| chr7 | 403621 | AAT | GAA | PF3D7_0709000 | crt | N75 | Asn75Glu | crt-Asn75Glu |
| chr7 | 404836 | C | G | PF3D7_0709000 | crt | Q271 | Gln271Glu | crt-Gln271Glu |
| chr7 | 403700 | G | T | PF3D7_0709000 | crt | C101 | Cys101Phe | crt-Cys101Phe |
| chr7 | 403620 | G | T | PF3D7_0709000 | crt | M74 | Met74Ile | crt-Met74Ile |
| chr7 | 405838 | G | T | PF3D7_0709000 | crt | R371 | Arg371Ile | crt-Arg371Ile |
| chr7 | 404010 | T | A | PF3D7_0709000 | crt | F145 | Phe145Ile | crt-Phe145Ile |
| chr7 | 403688 | A | T | PF3D7_0709000 | crt | H97 | His97Leu | crt-His97Leu |
| chr7 | 404407 | G | T | PF3D7_0709000 | crt | A220 | Ala220Ser | crt-Ala220Ser |
| chr7 | 403612 | T | A | PF3D7_0709000 | crt | C72 | Cys72Ser | crt-Cys72Ser |
| chr7 | 405362 | A | G | PF3D7_0709000 | crt | N326 | Asn326Ser | crt-Asn326Ser |
| chr7 | 405600 | T | C | PF3D7_0709000 | crt | I356 | Ile356Thr | crt-Ile356Thr |
| chr7 | 403625 | A | C | PF3D7_0709000 | crt | K76 | Lys76Thr | crt-Lys76Thr |
| chr7 | 403687 | C | T | PF3D7_0709000 | crt | H97 | His97Tyr | crt-His97Tyr |
| chr7 | 896660 | G | T | PF3D7_0720700 | pib7 | C1484 | Cys1484Phe | pib7-Cys1484Phe |
| chr8 | 549685 | G | C | PF3D7_0810800 | dhps | G437 | Gly437Ala | dhps-Gly437Ala |
| chr8 | 549681 | T | G | PF3D7_0810800 | dhps | S436 | Ser436Ala | dhps-Ser436Ala |
| chr8 | 549993 | A | G | PF3D7_0810800 | dhps | K540 | Lys540Glu | dhps-Lys540Glu |
| chr8 | 549682 | C | T | PF3D7_0810800 | dhps | S436 | Ser436Phe | dhps-Ser436Phe |
| chr8 | 549666 | A | G | PF3D7_0810800 | dhps | I431 | Ile431Val | dhps-Ile431Val |
| chr8 | 550117 | C | G | PF3D7_0810800 | dhps | A581 | Ala581Gly | dhps-Ala581Gly |
| chr8 | 550212 | G | T | PF3D7_0810800 | dhps | A613 | Ala613Ser | dhps-Ala613Ser |
| chr8 | 550212 | G | A | PF3D7_0810800 | dhps | A613 | Ala613Thr | dhps-Ala613Thr |
| chr10 | 950343 | C | A | PF3D7_1022600 | kelch10 | P623 | Pro623Thr | kelch10-Pro623Thr |

| | | | | | | | | |
|---|---|---|---|---|---|---|---|---|
| chr10 | 490720 | G | C | PF3D7_1012700 | pph | V1157 | Val1157Leu | pph-Val1157Leu |
| chr13 | 2504560 | A | G | PF3D7_1362500 | exo | E415 | Glu415Gly | exo-Glu415Gly |
| chr13 | 748395 | C | A | PF3D7_1318100 | fd | D193 | Asp193Tyr | fd-Asp193Tyr |
| chr13 | 1725521 | A | G | PF3D7_1343700 | k13 | Y493 | Tyr493His | k13-Tyr493His |
| chr13 | 1725570 | C | A | PF3D7_1343700 | k13 | M476 | Met476Ile | k13-Met476Ile |
| chr13 | 1725370 | A | G | PF3D7_1343700 | k13 | I543 | Ile543Thr | k13-Ile543Thr |
| chr13 | 1725382 | C | G | PF3D7_1343700 | k13 | R539 | Arg539Thr | k13-Arg539Thr |
| chr13 | 1725259 | C | T | PF3D7_1343700 | k13 | C580 | Cys580Tyr | k13-Cys580Tyr |
| chr13 | 2728402 | T | C | PF3D7_1368700 | mcp | N252 | Asn252Asp | mcp-Asn252Asp |
| chr13 | 958469 | G | A | PF3D7_1322700 | PF3D7-13 | T236 | Thr236Ile | PF3D7-1322700-Thr236Ile |
| chr14 | 2481070 | G | A | PF3D7_1460900.1 | arps10 | V127 | Val127Met | arps10-Val127Met |
| chr14 | 1956225 | G | A | PF3D7_1447900 | mdr2 | T484 | Thr484Ile | mdr2-Thr484Ile |
| chr14 | 2098642 | A | G | PF3D7-1451200 | PF3D7-14 | N71 | Asn71Asn | PF3D7-1451200-Asn71Asn |
| chrM | 4294 | A | G | mal_mito_3 | cytb | Y268 | Tyr268Cys | cytb-Tyr268Cys |
| chrM | 3890 | G | A | mal_mito_3 | cytb | M133 | Met133Ile | cytb-Met133Ile |
| chrM | 4341 | GT | AA | mal_mito_3 | cytb | V284 | Val284Lys | cytb-Val284Lys |
| chrM | 4293 | T | A | mal_mito_3 | cytb | Y268 | Tyr268Asn | cytb-Tyr268Asn |
| chrM | 4294 | A | C | mal_mito_3 | cytb | Y268 | Tyr268Ser | cytb-Tyr268Ser |

**Table S2. List of countries visited by travelers returning to France over 2016-2023**

| Visited country | n | % | rank | region |
|---|---|---|---|---|
| Côte d'Ivoire | 217 | 26.95652174 | 1 | WAF |
| Cameroon | 169 | 20.99378882 | 2 | CAF |
| Mali | 69 | 8.571428571 | 3 | WAF |
| Guinea | 58 | 7.204968944 | 4 | WAF |
| Republic of the Congo | 45 | 5.590062112 | 5 | CAF |
| Senegal | 37 | 4.596273292 | 6 | WAF |
| Central African Republic | 29 | 3.602484472 | 7 | CAF |
| Chad | 26 | 3.229813665 | 8 | CAF |
| Benin | 23 | 2.857142857 | 9 | WAF |
| Gabon | 22 | 2.732919255 | 10 | CAF |
| Burkina Faso | 17 | 2.111801242 | 11 | WAF |
| Togo | 17 | 2.111801242 | 12 | WAF |
| Nigeria | 12 | 1.49068323 | 13 | WAF |
| Niger | 11 | 1.366459627 | 14 | WAF |
| Democratic Republic of the Congo | 9 | 1.118012422 | 15 | CAF |
| Sierra Leone | 6 | 0.7453416149 | 16 | WAF |
| Comoros | 5 | 0.6211180124 | 17 | EAF |
| Angola | 3 | 0.3726708075 | 18 | CAF |
| Equatorial Guinea | 3 | 0.3726708075 | 19 | CAF |
| Ghana | 3 | 0.3726708075 | 20 | WAF |
| Madagascar | 3 | 0.3726708075 | 21 | EAF |
| Mozambique | 3 | 0.3726708075 | 22 | EAF |
| Africa without another indication | 2 | 0.248447205 | 23 | |
| Bangladesh | 1 | 0.1242236025 | 24 | SA |
| Burundi | 1 | 0.1242236025 | 25 | EAF |
| Egypt | 1 | 0.1242236025 | 26 | NAF |
| Kenya | 1 | 0.1242236025 | 27 | EAF |
| Liberia | 1 | 0.1242236025 | 28 | WAF |
| Malawi | 1 | 0.1242236025 | 29 | EAF |
| Morocco | 1 | 0.1242236025 | 30 | NAF |
| Mayotte | 1 | 0.1242236025 | 31 | EAF |
| Nauru | 1 | 0.1242236025 | 32 | OCE |
| No information | 1 | 0.1242236025 | 33 | |
| Uganda | 1 | 0.1242236025 | 34 | EAF |
| Rwanda | 1 | 0.1242236025 | 35 | EAF |
| Sudan | 1 | 0.1242236025 | 36 | EAF |
| Tanzania | 1 | 0.1242236025 | 37 | EAF |
| Thailand | 1 | 0.1242236025 | 38 | SEA |
| Zambia | 1 | 0.1242236025 | 39 | EAF |

CAF: Central Africa; EAF: East Africa; NAF: North Africa; OCE: Oceania; SA: South Asia; SEA: South East Asia; WAF: West Africa.

**Table S3. Frequency of isolates from West and Central Africa region in 2016-18 and 2019-23**

| region | 2016-18 (N=485) n (%) | 2019-23 (N=292) n (%) | *p-value** |
|---|---|---|---|
| CAF | 185 (38.1%) | 121 (41.4%) | 0.404 |
| WAF | 300 (61.9%) | 171 (58.6%) | |

CAF: Central Africa; WAF: West Africa.

*Significance of change in frequency of isolates in 2016-18 versus 2019-23 calculated by Chi-square test.

**Table S4. Drug susceptibility of control strain 3D7.**

| Drugs | N | $IC_{50}$ (nM) median [IQR] |
|---|---|---|
| Dihydroartemisinin | 69 | 1.48 [1.08, 2.02] |
| Lumefantrine | 83 | 18.7 [13.8, 33.9] |
| Mefloquine | 46 | 54.2 [28.0, 72.7] |
| Monodesethylamodiaquine | 65 | 21.6 [16.3, 25.6] |
| Chloroquine | 72 | 29.3 [24.1, 32.5] |
| Piperaquine | 78 | 24.2 [19.9, 28.2] |

**Table S5. Drug susceptibility in 2016-18 versus 2019-23**

| Drug | 2016-18 (N = 498) IC50 (nM) median [IQR] | 2019-23 (N = 307) IC50 (nM) median [IQR] | *p-value** |
|---|---|---|---|
| Dihydroartemisinin | 1.10 [0.73-1.64] | 1.14 [0.86-1.72] | 0.048 |
| Lumefantrine | 13.9 [8.42-21.7] | 23.0 [14.4-35.1] | 6.60E-16 |
| Mefloquine | 30.7 [19.9-45.1] | 28.2 [17.7-46.0] | 0.47 |
| Monodesethylamodiaquine | 20.3 [15.4-33.1] | 35.4 [21.2-51.1] | 6.60E-16 |
| Chloroquine | 27.7 [19.0-45.0] | 25.1 [17.3-38.9] | 0.048 |
| Piperaquine | 18.0 [14.2-22.4] | 20.5 [16.5-26.2] | 3.17E-06 |

IQR: Interquartile range. *Significance of change in susceptibility over time calculated by Mann-Whitney test. P-values were adjusted using Benjamini-Hochberg correction.

**Table S6. Drug susceptibility in 2016-18 versus 2019-23 by African region.**

| Drug | CAF | | | WAF | | |
|---|---|---|---|---|---|---|
| | 2016-18 (N=185) IC$_{50}$ (nM) median [IQR] | 2019-23 (N=121) IC$_{50}$ (nM) median [IQR] | *p-value** | 2016-18 (N=300) IC$_{50}$ (nM) median [IQR] | 2019-23 (N=171) IC$_{50}$ (nM) median [IQR] | *p-value** |
| DHA | 1.09 [0.75-1.63] | 1.28 [0.92-1.94] | 0.017 | 1.10 [0.71-1.63] | 1.10 [0.82-1.53] | 0.539 |
| LMF | 14.1 [7.89-22.2] | 23.0 [15.0-33.3] | 6.60E-16 | 13.9 [8.65-21.0] | 23.4 [14.6-36.1] | 6.60E-16 |
| MFQ | 31.1 [19.6-45.2] | 29.6 [20.6-45.6] | 0.926 | 30.5 [20.6-45.1] | 27.4 [17.0-49.0] | 0.610 |
| MDAQ | 19.7 [14.1-28.4] | 36.7 [22.7-50.6] | 6.60E-16 | 20.8 [16.4-34.8] | 34.7 [20.0-52.2] | 6.60E-16 |
| CQ | 27.3 [18.3-40.0] | 25.6 [17.3-35.2] | 0.274 | 27.8 [19.5-49.4] | 24.7 [17.4-44.3] | 0.194 |
| PPQ | 18.5 [15.7-24.2] | 21.9 [17.0-28.5] | 0.017 | 17.5 [13.6-21.1] | 19.8 [16.2-25.6] | 1.55E-06 |

DHA: Dihydroartemisinin; LMF: Lumefantrine; MFQ: Mefloquine; MDAQ: Monodesethylamodiaquine; CQ: Chloroquine- PPQ: Piperaquine.
CAF: Central Africa; WAF: West Africa. IQR: Interquartile range. *Significance of change in susceptibility over time calculated by Mann-Whitney test. P-values were adjusted using Benjamini-Hochberg correction.

**Table S7. Drug susceptibility by year**

| Drug | Year | N | IC50 (nM)- median [IQR] | Mann-Kendall tau [&] | *p-value* * |
|---|---|---|---|---|---|
| Dihydroartemisinin | 2016 | 165 | 1.10 [0.80- 1.60] | 0.33 | 0.469 |
| | 2017 | 166 | 1.16 [0.65- 2.62] | | |
| | 2018 | 167 | 1.05 [0.70- 1.50] | | |
| | 2019 | 100 | 0.94 [0.71- 1.24] | | |
| | 2020-21 | 85 | 1.28 [0.89- 1.74] | | |
| | 2022-23 | 122 | 1.42 [1.00- 2.21] | | |
| Lumefantrine | 2016 | 165 | 15.5 [7.87- 24.6] | 0.60 | 0.136 |
| | 2017 | 166 | 14.7 [9.76- 21.0] | | |
| | 2018 | 167 | 11.3 [7.45- 19.3] | | |
| | 2019 | 100 | 18.0 [11.8- 29.0] | | |
| | 2020-21 | 85 | 24.8 [16.8- 36.2] | | |
| | 2022-23 | 122 | 25.3 [15.6- 38.6] | | |
| Mefloquine | 2016 | 165 | 33.3 [22.8- 44.9] | -0.07 | 1.000 |
| | 2017 | 166 | 40.5 [24.7- 57.2] | | |
| | 2018 | 167 | 23.7 [15.9- 35.4] | | |
| | 2019 | 100 | 20.1 [13.6- 29.2] | | |
| | 2020-21 | 85 | 40.7 [23.4- 56.2] | | |
| | 2022-23 | 122 | 30.6 [21.4- 60.0] | | |
| Monodesethylamodiaquine | 2016 | 165 | 19.7 [13.1- 27.0] | 0.60 | 0.136 |
| | 2017 | 166 | 19.0 [15.9- 26.7] | | |
| | 2018 | 167 | 25.3 [17.9- 41.0] | | |
| | 2019 | 100 | 35.0 [22.8- 45.0] | | |
| | 2020-21 | 85 | 36.9 [23.7- 41.8] | | |
| | 2022-23 | 122 | 34.8 [18.2- 56.7] | | |
| Chloroquine | 2016 | 165 | 24.3 [17.1- 45.3] | 0.14 | 0.702 |
| | 2017 | 166 | 27.3 [19.5- 50.7] | | |
| | 2018 | 167 | 29.8 [20.3- 41.3] | | |
| | 2019 | 100 | 28.7 [18.3- 41.1] | | |
| | 2020-21 | 85 | 29.8 [21.0- 46.1] | | |
| | 2022-23 | 122 | 19.3 [14.6- 33.0] | | |
| Piperaquine | 2016 | 165 | 18.0 [14.2- 24.3] | 0.33 | 0.469 |
| | 2017 | 166 | 19.6 [16.8- 24.3] | | |
| | 2018 | 167 | 16.1 [11.9- 19.1] | | |
| | 2019 | 100 | 19.1 [13.5- 24.3] | | |
| | 2020-21 | 85 | 25.4 [19.3- 32.3] | | |
| | 2022-23 | 122 | 19.5 [16.7- 24.7] | | |

[&] Magnitude and direction of trend over time. IQR: Interquartile range.

*Significance of change in susceptibility over time by Mann-Kendall test.

**Table S8. Frequency of resistant isolates defined by cut-off$ in 2016-18 and 2019-23**

| **Drug** | **2016-18 (N=498) n (%)** | **2019-23 (N=307) n (%)** | ***p-value**** |
|---|---|---|---|
| Dihydroartemisinin | | | |
| R: >10nM | 1 (0.2%) | 0 (0%) | 1.00 |
| S: ≤10nM | 497 (99.8%) | 307 (100%) | |
| Lumefantrine | | | |
| R: >150nM | 0 (0%) | 3 (1.0%) | 0.11 |
| S: ≤150nM | 498 (100%) | 304 (99.0%) | |
| Mefloquine | | | |
| R: >30nM | 256 (51.4%) | 138 (45.0%) | 0.09 |
| S: ≤30nM | 242 (48.6%) | 169 (55.0%) | |
| Monodesethylamodiaquine | | | |
| R: >80nM | 12 (2.4%) | 11 (3.6%) | 0.45 |
| S: ≤80nM | 486 (97.6%) | 296 (96.4%) | |
| Chloroquine | | | |
| R: >100nM | 61 (12.2%) | 21 (6.8%) | 0.02 |
| S: ≤100nM | 437 (87.8%) | 286 (93.2%) | |
| Piperaquine | | | |
| R: >135nM | 1 (0.2%) | 0 (0%) | 1.00 |
| S: ≤135nM | 497 (99.8%) | 307 (100%) | |

R: resistant; S: sensitive. $ Cut-off as reported in Pascual et al. 2015 and Kaddouri et al. 2008. *Significance of change in frequency of resistant isolates in 2016-18 versus 2019-23 calculated by Chi-square test.

**References**:


-Pascual- Aurélie- Marilyn Madamet- Sébastien Briolant- Tiphaine Gaillard- Rémy Amalvict- Nicolas Benoit- Dominique Travers- Bruno Pradines- and French National Reference Centre for Imported Malaria Study Group. 2015. “Multinormal in Vitro Distribution of Plasmodium Falciparum Susceptibility to Piperaquine and Pyronaridine.” Malaria Journal 14 (February): 49.

-Kaddouri- Halima- Abdoulaye Djimdé- Souleymane Dama- Aly Kodio- Mamadou Tekete- Véronique Hubert- Aminatou Koné- et al. 2008. “Baseline in Vitro Efficacy of ACT Component Drugs on Plasmodium Falciparum Clinical Isolates from Mali.” International Journal for Parasitology 38 (7): 791–98.

**Table S9. Prevalence (%) of key mutations in the top ten countries visited by participants of this study from 2016-2023.**

| Gene | Mutation | Mali | Gabon | Ivory Coast | Republic of the Congo | Cameroon | Benin | Central African Republic | Senegal | Guinea | Chad |
|---|---|---|---|---|---|---|---|---|---|---|---|
| crt | Cys72Ser | 0.0 | 0.0 | 0.0 | 0.0 | 0.0 | 0.0 | 0.0 | 0.0 | 0.0 | 0.0 |
| crt | Met74Ile | 39.7 | 33.3 | 20.3 | 52.3 | 14.6 | 57.1 | 0.0 | 64.5 | 72.2 | 28.0 |
| crt | Asn75Glu | 39.7 | 33.3 | 20.3 | 52.3 | 14.6 | 57.1 | 0.0 | 64.5 | 72.2 | 28.0 |
| crt | Lys76Thr | 39.7 | 33.3 | 20.3 | 52.3 | 14.6 | 57.1 | 0.0 | 64.5 | 72.2 | 28.0 |
| crt | Thr93Ser | 0.0 | 0.0 | 0.0 | 0.0 | 0.0 | 0.0 | 0.0 | 0.0 | 0.0 | 0.0 |
| crt | His97Tyr | 0.0 | 0.0 | 0.0 | 0.0 | 0.6 | 0.0 | 0.0 | 0.0 | 0.0 | 0.0 |
| crt | Phe145Ile | 0.0 | 0.0 | 0.0 | 0.0 | 0.0 | 0.0 | 0.0 | 0.0 | 0.0 | 0.0 |
| crt | Ile218Phe | 0.0 | 0.0 | 0.6 | 0.0 | 2.3 | 0.0 | 0.0 | 0.0 | 0.0 | 0.0 |
| crt | Ala220Ser | 44.6 | 28.6 | 23.4 | 56.3 | 14.5 | 63.2 | 0.0 | 71.4 | 69.6 | 18.2 |
| crt | Gln271Glu | 43.9 | 35.7 | 23.9 | 45.2 | 12.8 | 57.9 | 0.0 | 62.1 | 70.2 | 20.0 |
| crt | Asn326Ser | 1.6 | 0.0 | 3.6 | 2.5 | 0.7 | 0.0 | 0.0 | 3.1 | 1.9 | 4.2 |
| crt | Met343Leu | 0.0 | 0.0 | 0.0 | 0.0 | 0.0 | 0.0 | 0.0 | 0.0 | 0.0 | 0.0 |
| crt | Gly353Val | 0.0 | 0.0 | 0.0 | 0.0 | 0.0 | 0.0 | 0.0 | 0.0 | 0.0 | 0.0 |
| crt | Ile356Thr | 28.6 | 35.3 | 8.1 | 29.4 | 11.7 | 45.0 | 0.0 | 43.3 | 64.0 | 12.0 |
| crt | Arg371Ile | 43.1 | 31.3 | 20.8 | 45.9 | 15.1 | 52.6 | 0.0 | 59.4 | 71.4 | 26.9 |
| dhfr-ts | Ala16Val | 0.0 | 0.0 | 0.0 | 0.0 | 0.0 | 0.0 | 0.0 | 0.0 | 0.0 | 0.0 |
| dhfr-ts | Asn51Ile | 81.5 | 100.0 | 79.2 | 100.0 | 98.0 | 100.0 | 95.8 | 87.9 | 92.2 | 96.0 |
| dhfr-ts | Cys59Arg | 84.6 | 100.0 | 86.5 | 92.9 | 99.3 | 100.0 | 95.8 | 93.9 | 94.1 | 92.0 |
| dhfr-ts | Ser108Asn | 83.1 | 100.0 | 87.9 | 100.0 | 98.7 | 100.0 | 100.0 | 90.9 | 96.0 | 100.0 |
| dhfr-ts | Ile164Leu | 0.0 | 0.0 | 0.0 | 0.0 | 0.0 | 0.0 | 0.0 | 0.0 | 0.0 | 4.2 |
| dhps | Ser436Ala | 64.5 | 25.0 | 64.0 | 18.9 | 45.6 | 42.9 | 52.4 | 30.0 | 58.0 | 76.0 |
| dhps | Ser436Phe | 0.0 | 0.0 | 2.7 | 0.0 | 0.0 | 0.0 | 0.0 | 0.0 | 0.0 | 0.0 |
| dhps | Ala437Gly | 51.6 | 87.5 | 53.8 | 89.2 | 68.0 | 71.4 | 61.9 | 70.0 | 58.0 | 28.0 |
| dhps | Lys540Glu | 3.2 | 5.3 | 1.0 | 18.4 | 3.9 | 0.0 | 24.0 | 6.1 | 10.0 | 4.0 |
| dhps | Ala581Gly | 1.5 | 5.0 | 5.0 | 14.6 | 12.6 | 19.0 | 0.0 | 0.0 | 1.9 | 19.2 |
| dhps | Ala613Ser | 14.1 | 0.0 | 19.8 | 5.0 | 16.5 | 26.3 | 0.0 | 3.1 | 18.5 | 15.4 |
| dhps | Ala613Thr | 0.0 | 0.0 | 0.0 | 0.0 | 0.0 | 0.0 | 0.0 | 0.0 | 0.0 | 0.0 |
| k13 | Met476Ile | 0.0 | 0.0 | 0.0 | 0.0 | 0.0 | 0.0 | 0.0 | 0.0 | 0.0 | 0.0 |
| k13 | Tyr493His | 1.6 | 0.0 | 0.0 | 0.0 | 0.0 | 0.0 | 0.0 | 0.0 | 0.0 | 0.0 |
| k13 | Arg539Thr | 0.0 | 0.0 | 0.0 | 0.0 | 0.0 | 0.0 | 0.0 | 0.0 | 0.0 | 0.0 |
| k13 | Ile543Thr | 0.0 | 0.0 | 0.5 | 0.0 | 0.0 | 0.0 | 0.0 | 0.0 | 0.0 | 0.0 |
| k13 | Arg561His | 0.0 | 0.0 | 0.0 | 0.0 | 0.0 | 0.0 | 0.0 | 0.0 | 0.0 | 0.0 |
| k13 | Ala578Ser | 0.0 | 0.0 | 0.5 | 0.0 | 3.3 | 0.0 | 0.0 | 0.0 | 3.8 | 0.0 |
| k13 | Cys580Tyr | 0.0 | 0.0 | 0.0 | 0.0 | 0.0 | 0.0 | 0.0 | 0.0 | 0.0 | 0.0 |
| k13 | Glu612Asp | 0.0 | 0.0 | 0.0 | 0.0 | 0.0 | 0.0 | 0.0 | 3.1 | 0.0 | 0.0 |
| k13 | Ala675Val | 0.0 | 0.0 | 0.0 | 0.0 | 0.0 | 0.0 | 0.0 | 0.0 | 0.0 | 0.0 |
| mdr1 | Asn86Tyr | 10.6 | 0.0 | 7.4 | 11.9 | 15.2 | 4.8 | 0.0 | 23.5 | 17.3 | 3.8 |
| mdr1 | Tyr184Phe | 75.4 | 61.1 | 70.4 | 63.6 | 70.3 | 65.2 | 80.8 | 67.6 | 76.8 | 69.2 |
| mdr1 | Ser1034Cys | 0.0 | 0.0 | 0.0 | 0.0 | 1.2 | 0.0 | 0.0 | 0.0 | 0.0 | 0.0 |
| mdr1 | Asn1042Asp | 0.0 | 0.0 | 0.0 | 0.0 | 0.0 | 0.0 | 0.0 | 0.0 | 0.0 | 0.0 |
| mdr1 | Asp1246Tyr | 0.0 | 0.0 | 1.1 | 2.6 | 0.6 | 4.3 | 0.0 | 0.0 | 0.0 | 0.0 |

**Table S10. Frequency distribution of *pfcrt* haplotypes in the ten top countries visited by participants of this study**

| haplotype | Total (N=487) | Ivory Coast (N=128) | Cameroon (N=104) | Mali (N=46) | Guinea (N=36) | Republic of the Congo (N=23) | Senegal (N=19) | Chad (N=18) | Central African Republic (N=14) | Gabon (N=13) | Benin (N=12) |
|---|---|---|---|---|---|---|---|---|---|---|---|
| MNKAQNIR (Wild-type) | 374 (76.8%) | 108 (84.4%) | 96 (92.3%) | 30 (65.2%) | 12 (33.3%) | 17 (73.9%) | 8 (42.1%) | 15 (83.3%) | 14 (100%) | 9 (69.2%) | 5 (41.7%) |
| **IETSE**NI**I** (GB4) | 30 (6.2%) | 8 (6.3%) | 1 (1.0%) | 5 (10.9%) | 4 (11.1%) | 4 (17.4%) | 1 (5.3%) | 1 (5.6%) | 0 (0%) | 0 (0%) | 0 (0%) |
| **IETSE**N**TI** (Cam783) | 75 (15.4%) | 8 (6.3%) | 7 (6.7%) | 10 (21.7%) | 20 (55.6%) | 2 (8.7%) | 9 (47.4%) | 1 (5.6%) | 0 (0%) | 4 (30.8%) | 7 (58.3%) |
| **IETSES**I**I** (FCB) | 3 (0.6%) | 0 (0%) | 0 (0%) | 0 (0%) | 0 (0%) | 0 (0%) | 1 (5.3%) | 1 (5.6%) | 0 (0%) | 0 (0%) | 0 (0%) |
| MNK**SE**NI**I** | 3 (0.6%) | 2 (1.6%) | 0 (0%) | 1 (2.2%) | 0 (0%) | 0 (0%) | 0 (0%) | 0 (0%) | 0 (0%) | 0 (0%) | 0 (0%) |
| MNKA**E**NIR | 1 (0.2%) | 1 (0.8%) | 0 (0%) | 0 (0%) | 0 (0%) | 0 (0%) | 0 (0%) | 0 (0%) | 0 (0%) | 0 (0%) | 0 (0%) |
| MNK**S**QNIR | 1 (0.2%) | 1 (0.8%) | 0 (0%) | 0 (0%) | 0 (0%) | 0 (0%) | 0 (0%) | 0 (0%) | 0 (0%) | 0 (0%) | 0 (0%) |

*Pfcrt* haplotypes built with amino acid positions 74- 75- 76- 220- 271- 326- 356 and 371. Letters in bold red represent the amino acid mutations.

**Table S11. Prevalence of ART-R background mutations**

| Gene | Mutation | Total (n) | Mutant (n) | Prevalence (%) |
|---|---|---|---|---|
| crt | Asn326Ser | 710 | 14 | 1.97 |
| crt | Ile356Thr | 686 | 133 | 19.39 |
| fd | Asp193Tyr | 737 | 2 | 0.27 |
| mdr2 | Thr484Ile | 628 | 0 | 0.00 |
| pib7 | Cys1484Phe | 642 | 1 | 0.16 |
| pph | Val1157Leu | 677 | 0 | 0.00 |

**Table S12. Prevalence (%) of key mutations by year in all isolates**

| Gene | Mutation | 2016 (%) | 2017 (%) | 2018 (%) | 2019 (%) | 2020-21 (%) | 2022-23 (%) |
|---|---|---|---|---|---|---|---|
| crt | Cys72Ser | 0.00 | 0.00 | 0.00 | 0.00 | 0.00 | 0.00 |
| crt | Met74Ile | 36.30 | 32.05 | 30.25 | 30.56 | 22.67 | 22.61 |
| crt | Asn75Glu | 36.30 | 32.05 | 30.25 | 30.56 | 22.67 | 22.61 |
| crt | Lys76Thr | 36.30 | 32.05 | 30.25 | 30.56 | 22.67 | 22.61 |
| crt | Thr93Ser | 0.00 | 0.00 | 0.00 | 0.00 | 0.00 | 0.00 |
| crt | His97Tyr | 0.68 | 0.00 | 0.00 | 0.00 | 0.00 | 0.00 |
| crt | Phe145Ile | 0.00 | 0.00 | 0.00 | 0.00 | 0.00 | 0.00 |
| crt | Ile218Phe | 0.00 | 1.40 | 0.70 | 1.61 | 0.00 | 0.00 |
| crt | Ala220Ser | 35.65 | 32.17 | 30.77 | 35.48 | 25.93 | 23.30 |
| crt | Gln271Glu | 35.45 | 34.09 | 33.11 | 27.59 | 20.75 | 23.36 |
| crt | Asn326Ser | 4.26 | 0.00 | 1.85 | 1.37 | 1.43 | 2.65 |
| crt | Met343Leu | 0.00 | 0.00 | 0.00 | 0.00 | 0.00 | 0.00 |
| crt | Gly353Val | 0.00 | 0.00 | 0.00 | 0.00 | 0.00 | 0.00 |
| crt | Ile356Thr | 17.46 | 22.97 | 22.44 | 20.59 | 17.57 | 13.16 |
| crt | Arg371Ile | 34.11 | 31.94 | 30.82 | 28.36 | 23.19 | 22.81 |
| dhfr-ts | Ala16Val | 0.00 | 0.00 | 0.00 | 0.00 | 0.00 | 0.00 |
| dhfr-ts | Asn51Ile | 88.36 | 85.90 | 91.93 | 85.14 | 94.52 | 91.15 |
| dhfr-ts | Cys59Arg | 89.04 | 91.03 | 95.65 | 90.54 | 91.78 | 95.58 |
| dhfr-ts | Ser108Asn | 90.97 | 92.26 | 96.88 | 91.30 | 94.37 | 94.83 |
| dhfr-ts | Ile164Leu | 0.00 | 0.00 | 0.62 | 0.00 | 0.00 | 0.00 |
| dhps | Ser436Ala | 46.04 | 53.25 | 52.83 | 43.28 | 55.38 | 52.78 |
| dhps | Ser436Phe | 0.72 | 0.65 | 1.89 | 0.00 | 0.00 | 0.93 |
| dhps | Ala437Gly | 64.03 | 59.09 | 58.49 | 65.67 | 55.38 | 66.67 |
| dhps | Lys540Glu | 5.67 | 8.44 | 4.97 | 4.17 | 5.19 | 8.70 |
| dhps | Ala581Gly | 4.93 | 10.56 | 8.54 | 7.41 | 11.39 | 5.79 |
| dhps | Ala613Ser | 8.57 | 17.61 | 19.51 | 11.84 | 19.74 | 12.50 |
| dhps | Ala613Thr | 0.00 | 0.00 | 0.00 | 0.00 | 0.00 | 0.00 |
| k13 | Met476Ile | 0.00 | 0.00 | 0.00 | 0.00 | 0.00 | 0.00 |
| k13 | Tyr493His | 0.71 | 0.00 | 0.00 | 0.00 | 0.00 | 0.00 |
| k13 | Arg539Thr | 0.00 | 0.00 | 0.00 | 0.00 | 0.00 | 0.00 |
| k13 | Ile543Thr | 0.00 | 0.00 | 0.61 | 0.00 | 0.00 | 0.00 |
| k13 | Arg561His | 0.00 | 0.00 | 0.00 | 0.00 | 0.00 | 0.00 |
| k13 | Ala578Ser | 2.78 | 1.97 | 0.00 | 0.00 | 0.00 | 0.91 |
| k13 | Cys580Tyr | 0.00 | 0.00 | 0.00 | 0.00 | 0.00 | 0.00 |
| k13 | Glu612Asp | 0.00 | 0.70 | 0.00 | 0.00 | 0.00 | 0.00 |
| k13 | Ala675Val | 0.00 | 0.00 | 0.00 | 1.39 | 1.30 | 0.00 |
| mdr1 | Asn86Tyr | 13.61 | 14.91 | 14.63 | 5.13 | 3.80 | 5.98 |
| mdr1 | Tyr184Phe | 73.58 | 70.55 | 75.61 | 71.95 | 62.96 | 65.00 |
| mdr1 | Ser1034Cys | 0.00 | 0.61 | 0.61 | 0.00 | 0.00 | 0.00 |
| mdr1 | Asn1042Asp | 0.00 | 0.00 | 0.00 | 0.00 | 0.00 | 0.00 |
| mdr1 | Asp1246Tyr | 0.00 | 1.31 | 0.62 | 2.82 | 0.00 | 0.88 |

**Table S13. Full list of mutations associated with $IC_{50}$ identified by targeted genome association tests**

| Drug | Chr | Gene | Mutation | p-value* |
|---|---|---|---|---|
| | **7** | **crt** | **Arg371Ile** | **1.04E-06** |
| | **7** | **crt** | **Gln271Glu** | **1.18E-06** |
| | **7** | **crt** | **Ala220Ser** | **2.09E-06** |
| | **7** | **crt** | **Asn75Glu** | **8.52E-06** |
| | **7** | **crt** | **Lys76Thr** | **8.52E-06** |
| | **7** | **crt** | **Met74Ile** | **8.52E-06** |
| | 7 | crt | Ile356Thr | 0.000174962 |
| | 1 | Pfubp1 | Lys906Asn | 0.00028029 |
| | 1 | Pfubp1 | Gly767Asp | 0.000288432 |
| | 5 | mdr1 | Asn86Tyr | 0.000346631 |
| | 12 | pfmrp2 | Ala1643Val | 0.001336461 |
| | 2 | PF3D7-0218600 | Lys650Asn | 0.001505604 |
| | 6 | PF3D7-0613800 | Tyr2413Asn | 0.001769895 |
| | 1 | Pfubp1 | Asp41Gly | 0.002191417 |
| | 13 | k13 | Arg255Lys | 0.005262165 |
| | 1 | Pfubp1 | Glu1528Asp | 0.005736937 |
| Dihydroartemisinin | 1 | Pfubp1 | Asn300Lys | 0.007144591 |
| | 14 | PM3 | Lys151Asn | 0.009843502 |
| | 5 | pfpi4k | Asn878Asp | 0.011502325 |
| | 7 | pfpare | Met261Ile | 0.013885198 |
| | 1 | Pfubp1 | Asn917Lys | 0.015889482 |
| | 1 | Pfubp1 | Asp777Gly | 0.01812707 |
| | 6 | PF3D7-0613800 | Tyr1944Asn | 0.018700233 |
| | 3 | pfabcI3 | Pro2965Arg | 0.025149814 |
| | 1 | atp6 | Leu402Val | 0.028539667 |
| | 2 | PF3D7-0218600 | Ala1441Thr | 0.029810326 |
| | 6 | PF3D7-0613800 | Gly2154Asp | 0.030017952 |
| | 6 | PF3D7-0613800 | Asp1149Asn | 0.039883334 |
| | 6 | PF3D7-0613800 | Gly1150Asn | 0.039883334 |
| | 6 | PF3D7-0613800 | Ile2934Leu | 0.040192265 |
| | 5 | mdr1 | Asp650Asn | 0.044090663 |
| | 1 | Pfubp1 | Glu1922Lys | 0.04579986 |
| | 1 | Pfubp1 | Lys1921Asn | 0.04579986 |
| | **7** | **crt** | **Asn75Glu** | **3.82E-05** |
| | **7** | **crt** | **Lys76Thr** | **3.82E-05** |
| | **7** | **crt** | **Met74Ile** | **3.82E-05** |
| | **5** | **mdr1** | **Asn86Tyr** | **6.66E-05** |
| | 7 | crt | Gln271Glu | 0.000215645 |
| | 7 | crt | Ala220Ser | 0.000415635 |
| | 2 | PF3D7-0218600 | Lys1689Ile | 0.000516778 |
| | 6 | PF3D7-0613800 | Lys221Gln | 0.00120823 |
| | 7 | crt | Arg371Ile | 0.001255971 |
| | 5 | mdr1 | Asn652Asp | 0.003673781 |
| | 7 | crt | Ile356Thr | 0.004607846 |
| Lumefantrine | 5 | mdr1 | Asp650Asn | 0.005730007 |
| | 1 | Pfubp1 | His3083Arg | 0.008863005 |
| | 6 | PF3D7-0613800 | Gln245Glu | 0.008886072 |
| | 6 | PF3D7-0613800 | Glu2160Gln | 0.013079529 |
| | 6 | Sec14 | Asn615Asp | 0.015390987 |
| | 12 | pfmrp2 | Gly623Asp | 0.020454366 |
| | 1 | Pfubp1 | Tyr1462His | 0.026162996 |
| | 8 | dhps | Ala437Gly | 0.031749515 |
| | 12 | Pfap2mu | Asn233Lys | 0.032426901 |
| | 2 | PF3D7-0218600 | Asn493Ser | 0.033533692 |
| | 1 | Pfmrp1 | Asn325Ser | 0.034403357 |
| | 14 | PM2 | Leu321Val | 0.034821752 |

| | | | | |
|---|---|---|---|---|
| | 12 | Pfatp4 | Asp1116Gly | 0.038554051 |
| | 1 | Pfubp1 | Asn1706Lys | 0.039811933 |
| | 1 | Pfubp1 | Asn157Lys | 0.041772223 |
| | 6 | PF3D7-0613800 | Lys1599Asn | 0.042366785 |
| | 6 | PF3D7-0613800 | Ala235Glu | 0.045343999 |
| | 2 | PF3D7-0218600 | Asn1556Ile | 0.049629144 |
| Mefloquine | **5** | **mdr1** | **Asn86Tyr** | **5.82E-07** |
| | 7 | crt | Ile356Thr | 0.00021839 |
| | 7 | crt | Ala220Ser | 0.000223342 |
| | 7 | crt | Asn75Glu | 0.000527672 |
| | 7 | crt | Lys76Thr | 0.000527672 |
| | 7 | crt | Met74Ile | 0.000527672 |
| | 5 | mdr1 | Asp650Asn | 0.000820005 |
| | 7 | crt | Gln271Glu | 0.001018982 |
| | 5 | mdr1 | Asn652Asp | 0.001502041 |
| | 6 | PF3D7-0613800 | Leu1663Phe | 0.002848381 |
| | 6 | PF3D7-0613800 | Glu700Ala | 0.002890115 |
| | 3 | pfabcI3 | Ser2728Leu | 0.003604245 |
| | 12 | pfaat2 | Gln608Lys | 0.00383045 |
| | 7 | crt | Arg371Ile | 0.005148505 |
| | 6 | PF3D7-0613800 | Tyr2743His | 0.008731524 |
| | 12 | pfmrp2 | Ala1643Val | 0.010417558 |
| | 2 | PF3D7-0218600 | Lys1689Ile | 0.010681292 |
| | 8 | dhps | Ser436Ala | 0.014688803 |
| | 2 | PF3D7-0218600 | Asp1550Ala | 0.019666369 |
| | 14 | PM1 | Thr24Ile | 0.022571106 |
| | 12 | pfaat2 | Arg625Lys | 0.031921953 |
| | 6 | PF3D7-0613800 | Lys2744Asn | 0.032689631 |
| | 3 | pfabcI3 | Ser2966Ala | 0.036603562 |
| | 3 | pfabcI3 | Pro2965Arg | 0.037606663 |
| | 12 | pfaat2 | Thr605Lys | 0.043115073 |
| | 1 | Pfubp1 | Tyr297Phe | 0.04332278 |
| | 12 | pfaat2 | Thr738Lys | 0.043958422 |
| | 12 | Pfatp4 | Gly1128Arg | 0.044843752 |
| | 1 | Pfubp1 | Asn1518Tyr | 0.04672036 |
| | 1 | Pfubp1 | Glu1519Asp | 0.04672036 |
| | 1 | Pfubp1 | Asp1525Glu | 0.047348731 |
| | 1 | Pfubp1 | Lys1705Asn | 0.04772637 |
| Monodesethylamodiaquine | **7** | **crt** | **Arg371Ile** | **1.28E-08** |
| | **7** | **crt** | **Asn75Glu** | **4.61E-08** |
| | **7** | **crt** | **Lys76Thr** | **4.61E-08** |
| | **7** | **crt** | **Met74Ile** | **4.61E-08** |
| | **7** | **crt** | **Ile356Thr** | **1.09E-07** |
| | **7** | **crt** | **Gln271Glu** | **1.51E-06** |
| | **7** | **crt** | **Ala220Ser** | **2.11E-06** |
| | 12 | pfmrp2 | Lys1842Ile | 0.001200511 |
| | 1 | Pfubp1 | Asn2813Lys | 0.002232132 |
| | 6 | PF3D7-0613800 | Glu2160Gln | 0.002256977 |
| | 1 | Pfubp1 | Gly2814Cys | 0.00273012 |
| | 1 | Pfubp1 | Asn769Lys | 0.006537341 |
| | 7 | pfpare | Lys322Arg | 0.011917587 |
| | 1 | Pfubp1 | Ser161Tyr | 0.014642815 |
| | 8 | dhps | Lys540Glu | 0.014960209 |
| | 6 | PF3D7-0613800 | Lys1131Asn | 0.015708988 |
| | 6 | PF3D7-0613800 | Asn3203Asp | 0.016091837 |
| | 1 | atp6 | Asn569Lys | 0.01630003 |
| | 13 | PF3D7-1322700 | Gly253Glu | 0.019192227 |
| | 14 | pfeef2 | Asn574His | 0.022295654 |
| | 1 | Pfubp1 | Asn1704Lys | 0.022678096 |

| | | | | |
|---|---|---|---|---|
| | 8 | dhps | Ala437Gly | 0.023575814 |
| | 2 | PF3D7-0218600 | Lys1689Ile | 0.023815834 |
| | 1 | atp6 | Gly639Asp | 0.029929163 |
| | 6 | PF3D7-0613800 | Asn1522His | 0.034322861 |
| | 12 | pfmrp2 | Ser734Asn | 0.034788461 |
| | 3 | pfabcI3 | Leu1830Ser | 0.035725504 |
| | 14 | PM1 | Thr347Ala | 0.039143878 |
| | 1 | Pfubp1 | Lys2809Asn | 0.041676131 |
| | 6 | PF3D7-0613800 | Asp1149Asn | 0.041996569 |
| | 6 | PF3D7-0613800 | Gly1150Asn | 0.041996569 |
| | 6 | PF3D7-0613800 | Arg1034Cys | 0.048782884 |
| Chloroquine | **7** | **crt** | **Arg371Ile** | **3.19E-50** |
| | **7** | **crt** | **Asn75Glu** | **1.48E-49** |
| | **7** | **crt** | **Lys76Thr** | **1.48E-49** |
| | **7** | **crt** | **Met74Ile** | **1.48E-49** |
| | **7** | **crt** | **Ala220Ser** | **5.00E-40** |
| | **7** | **crt** | **Ile356Thr** | **3.88E-39** |
| | **7** | **crt** | **Gln271Glu** | **1.60E-38** |
| | **1** | **Pfubp1** | **Asn1704Lys** | **2.19E-05** |
| | **1** | **Pfubp1** | **Lys1705Asn** | **0.000119618** |
| | 6 | pfaat1 | Ser258Leu | 0.001868487 |
| | 12 | pfaat2 | Val501Leu | 0.00243588 |
| | 1 | Pfubp1 | Arg2238Lys | 0.006096389 |
| | 7 | pfpare | Lys322Arg | 0.007980251 |
| | 12 | pfmrp2 | Lys1842Ile | 0.009830898 |
| | 5 | mdr1 | Asn86Tyr | 0.010637034 |
| | 12 | pfaat2 | Arg625Lys | 0.011126437 |
| | 3 | pfabcI3 | Ser2966Ala | 0.020627569 |
| | 5 | pfpi4k | Asn877Asp | 0.022979866 |
| | 1 | Pfubp1 | Asn520Lys | 0.023790564 |
| | 1 | Pfubp1 | Ser3112Asn | 0.029423528 |
| | 12 | pfmrp2 | His282Arg | 0.029899145 |
| | 7 | pfpare | Val349Ile | 0.03285974 |
| | 12 | pfaat2 | Glu810Gln | 0.036551102 |
| | 6 | PF3D7-0613800 | Lys1131Asn | 0.037143947 |
| | 6 | PF3D7-0613800 | Val229Glu | 0.037489827 |
| | 6 | PF3D7-0613800 | Asp230Gly | 0.044558238 |
| | 1 | Pfubp1 | Lys1193Thr | 0.046757001 |
| | 1 | Pfubp1 | Asp465Gly | 0.047411034 |
| Piperaquine | 7 | crt | Ala220Ser | 0.000231088 |
| | 7 | crt | Gln271Glu | 0.002285497 |
| | 6 | PF3D7-0613800 | Glu1946Asp | 0.004204042 |
| | 7 | pfpare | Met261Ile | 0.004453907 |
| | 6 | PF3D7-0613800 | Tyr1944Asn | 0.004838739 |
| | 1 | Pfubp1 | Gly767Asp | 0.012073952 |
| | 6 | PF3D7-0613800 | Asp1945Gly | 0.013467259 |
| | 1 | Pfubp1 | Cys2818Gly | 0.016759695 |
| | 1 | Pfubp1 | Lys2817Asn | 0.016759695 |
| | 6 | PF3D7-0613800 | Asp2635Val | 0.027765438 |
| | 6 | PF3D7-0613800 | Lys221Gln | 0.029913968 |
| | 1 | Pfubp1 | Lys895Asn | 0.032888657 |
| | 4 | dhfr | ts-Cys59Arg | 0.03321993 |
| | 2 | PF3D7-0218600 | Lys1689Ile | 0.033220959 |
| | 12 | pfaat2 | Ser614Arg | 0.034806383 |
| | 3 | pfcarl | Asp338Glu | 0.03576813 |
| | 7 | crt | Asn75Glu | 0.038632802 |
| | 7 | crt | Lys76Thr | 0.038632802 |
| | 7 | crt | Met74Ile | 0.038632802 |
| | 12 | Pfatp4 | Leu1124Arg | 0.043156704 |

| | | | | |
|---|---|---|---|---|
| | 2 | PF3D7-0218600 | Arg1442Cys | 0.044476293 |
| | 6 | PF3D7-0613800 | Thr2636Met | 0.049787867 |

Chr: Chromosome

* The models were adjusted by complexity of infection, year and the visited country of the imported case. Mutations highlighted in bold are the mutations that survived the Bonferroni correction.

**Table S14. Genomic loci associated with IC50 identified by targeted genome association tests**

| Initial TGAS | *P-value** | *P-value* (CRT K76T)* | *P-value* (CRT K76T and CRT I356T)* | *P-value* (MDR1 N86Y)* |
|---|---|---|---|---|
| **Dihydroartemisinin** | | | | |
| crt.Arg371Ile | 1.04E-06 | | | 1.23E-06 |
| crt.Gln271Glu | 1.18E-06 | | | 1.34E-06 |
| crt.Ala220Ser | 2.09E-06 | | | 1.19E-06 |
| crt.Asn75Glu | 8.52E-06 | | | 1.06E-05 |
| crt.Lys76Thr | 8.52E-06 | | | 1.06E-05 |
| crt.Met74Ile | 8.52E-06 | | | 1.06E-05 |
| **Lumefantrine** | | | | |
| crt.Asn75Glu | 3.82E-05 | | | 5.89E-05 |
| crt.Lys76Thr | 3.82E-05 | | | 5.89E-05 |
| crt.Met74Ile | 3.82E-05 | | | 5.89E-05 |
| mdr1.Asn86Tyr | 6.66E-05 | 0.000123392 | 9.67E-05 | |
| **Mefloquine** | | | | |
| mdr1.Asn86Ty | 5.82E-07 | 1.41E-05 | 1.92E-06 | |
| **Monodesethylamodiaquine** | | | | |
| crt.Arg371Ile | 1.28E-08 | | | 1.15E-08 |
| crt.Asn75Glu | 4.61E-08 | | | 4.23E-08 |
| crt.Lys76Thr | 4.61E-08 | | | 4.23E-08 |
| crt.Met74Ile | 4.61E-08 | | | 4.23E-08 |
| crt.Ile356Thr | 1.09E-07 | | | 1.12E-07 |
| crt.Gln271Glu | 1.51E-06 | | | 1.31E-06 |
| crt.Ala220Ser | 2.11E-06 | | | 1.97E-06 |
| **Chloroquine** | | | | |
| crt.Arg371Ile | 3.19E-50 | | | 4.58E-51 |
| crt.Asn75Glu | 1.48E-49 | 6.52E-07 | 3.36E-08 | 1.36E-49 |
| crt.Lys76Thr | 1.48E-49 | | | 1.36E-49 |
| crt.Met74Ile | 1.48E-49 | 6.52E-07 | 3.36E-08 | 1.36E-49 |
| crt.Ala220Ser | 5.00E-40 | | | 1.08E-40 |
| crt.Ile356Thr | 3.88E-39 | | | 6.90E-40 |
| crt.Gln271Glu | 1.60E-38 | | | 2.35E-39 |
| Pfubp1.Asn1704Lys | 2.19E-05 | | | 5.75E-05 |
| Pfubp1.Lys1705Asn | 1.20E-04 | | | |

TGAS: Targeted genome association test.

* The models were adjusted by complexity of infection- year and the visited country of the imported case.

*(CRT K76T): In this model- the presence of CRT (K76T ) SNP was added as an additional covariate.

*(CRT K76T and CRT I356T): In this model the presence of CRT (K76T ) and CRT (I356T) SNPs were added as additional covariates.

*(MDR1 N86Y): In this model the presence of MDR1 (N86Y) SNP was added as an additional covariate.

**Table S15. Distribution of $IC_{50}$ according to associated SNPs**

| Drug | Chr | Gene description | Mutation | genotype | n | median IC50 (Q1-Q3) | *P-value** |
|---|---|---|---|---|---|---|---|
| DHA | 7 | chloroquine resistance transporter (*pfcrt*) | Met74Ile | Wild type | 509 | 1.25 (0.87-1.91) | 8.52E-06 |
| | | | Met74Ile | Mutant | 217 | 0.92 (0.71-1.38) | |
| | | | Asn75Glu | Wild type | 509 | 1.25 (0.87-1.91) | 8.52E-06 |
| | | | Asn75Glu | Mutant | 217 | 0.92 (0.71-1.38) | |
| | | | Lys76Thr | Wild type | 509 | 1.2 5(0.87-1.91) | 8.52E-06 |
| | | | Lys76Thr | Mutant | 217 | 0.9 2(0.71-1.38) | |
| | | | Ala220Ser | Wild type | 429 | 1.26 (0.86-1.97) | 2.09E-06 |
| | | | Ala220Ser | Mutant | 191 | 0.9 2(0.71-1.38) | |
| | | | Gln271Glu | Wild type | 423 | 1.2 2(0.86-1.94) | 1.18E-06 |
| | | | Gln271Glu | Mutant | 185 | 0.9 3(0.66-1.37) | |
| | | | Arg371Ile | Wild type | 482 | 1.26 (0.87-1.94) | 1.04E-06 |
| | | | Arg371Ile | Mutant | 200 | 0.94 (0.69-1.4) | |
| LMF | 5 | multidrug resistance 1 (*pfmdr1*) | Asn86Tyr | Wild type | 664 | 17.85 (10.99-28.78) | 6.66E-05 |
| | | | Asn86Tyr | Mutant | 82 | 8.28 (4.21-15.89) | |
| | 7 | chloroquine resistance transporter (*pfcrt*) | Met74Ile | Wild type | 509 | 18.66 (11.69-30.01) | 3.82E-05 |
| | | | Met74Ile | Mutant | 217 | 12.56 (7.2-21.61) | |
| | | | Asn75Glu | Wild type | 509 | 18.66 (11.69-30.01) | 3.82E-05 |
| | | | Asn75Glu | Mutant | 217 | 12.56 (7.2-21.61) | |
| | | | Lys76Thr | Wild type | 509 | 18.66 (11.69-30.01) | 3.82E-05 |
| | | | Lys76Thr | Mutant | 217 | 12.56 (7.2-21.61) | |
| MFQ | 5 | multidrug resistance 1 (*pfmdr1*) | Asn86Tyr | Wild type | 664 | 31.89 (20.93-47.62) | 5.82E-07 |
| | | | Asn86Tyr | Mutant | 82 | 16.17 (10.94-27.84) | |
| MDAQ | 7 | chloroquine resistance transporter (*pfcrt*) | Met74Ile | Wild type | 509 | 21.02 (16.09-36.32) | 4.61E-08 |
| | | | Met74Ile | Mutant | 217 | 33.28 (20.7-48.39) | |
| | | | Asn75Glu | Wild type | 509 | 21.02 (16.09-36.32) | 4.61E-08 |
| | | | Asn75Glu | Mutant | 217 | 33.28 (20.7-48.39) | |
| | | | Lys76Thr | Wild type | 509 | 21.02 (16.09-36.32) | 4.61E-08 |
| | | | Lys76Thr | Mutant | 217 | 33.28 (20.7-48.39) | |
| | | | Arg371Ile | Wild type | 482 | 21.05 (15.94-36.22) | 1.28E-08 |
| | | | Arg371Ile | Mutant | 200 | 35.23 (22.13-50.32) | |
| | | | Ile356Thr | Wild type | 553 | 21.87 (16.77-37.57) | 1.09E-07 |
| | | | Ile356Thr | Mutant | 133 | 36.42 (23.25-55.32) | |
| | | | Ala220Ser | Wild type | 429 | 21.08 (15.92-35.9) | 2.11E-06 |
| | | | Ala220Ser | Mutant | 191 | 33.74 (21.18-44.58) | |
| | | | Gln271Glu | Wild type | 423 | 20.93 (15.99-35.96) | 1.51E-06 |
| | | | Gln271Glu | Mutant | 185 | 33.45 (21.28-48.05) | |
| CQ | 7 | chloroquine resistance transporter (*pfcrt*) | Arg371Ile | Wild type | 482 | 22.9 (16.34-30.46) | 3.19E-50 |
| | | | Arg371Ile | Mutant | 200 | 68.58 (34.84-130.6) | |
| | | | Met74Ile | Wild type | 509 | 22.82 (16.31-30.1) | 1.48E-49 |
| | | | Met74Ile | Mutant | 217 | 66.88 (33.15-125.35) | |
| | | | Asn75Glu | Wild type | 509 | 22.82 (16.31-30.1) | 1.48E-49 |

| | | | | | | |
|---|---|---|---|---|---|---|
| | | Asn75Glu | Mutant | 217 | 66.88 (33.15-125.35) | |
| | | Lys76Thr | Wild type | 509 | 22.82 (16.31-30.1) | 1.48E-49 |
| | | Lys76Thr | Mutant | 217 | 66.88 (33.15-125.35) | |
| | | Ala220Ser | Wild type | 429 | 22.61 (16.42-30.93) | 5.00E-40 |
| | | Ala220Ser | Mutant | 191 | 66.88 (32.6-128.03) | |
| | | Ile356Thr | Wild type | 553 | 24.2 (16.96-32.48) | 3.88E-39 |
| | | Ile356Thr | Mutant | 133 | 72.09 (37.44-148.29) | |
| | | Gln271Glu | Wild type | 423 | 23.32 (16.37-30.49) | 1.60E-38 |
| | | Gln271Glu | Mutant | 185 | 66.13 (33.15-125.09) | |
| | | Asn1704Lys | Wild type | 631 | 26.51 (17.54-38.79) | 2.19E-05 |
| 1 | ubiquitin-binding protein-1 (*pfubp-1*) | Asn1704Lys | Mutant | 54 | 32.3 (23.05-77.38) | |
| | | Lys1705Asn | Wild type | 617 | 26.51 (17.75-38.71) | 1.20E-04 |
| | | Lys1705Asn | Mutant | 68 | 32 (21.86-74.58) | |

DHA: Dihydroartemisinin; LMF: Lumefantrine; MFQ: Mefloquine; MDAQ: Monodesethylamodiaquine; CQ: Chloroquine
Chr: Chromosome; n : number of individuals; IC50: half-maximal inhibitory concentration in nM
Q1: first quartile; Q3: third quartile; *IC50 median were compared by Mann-Whitney test.

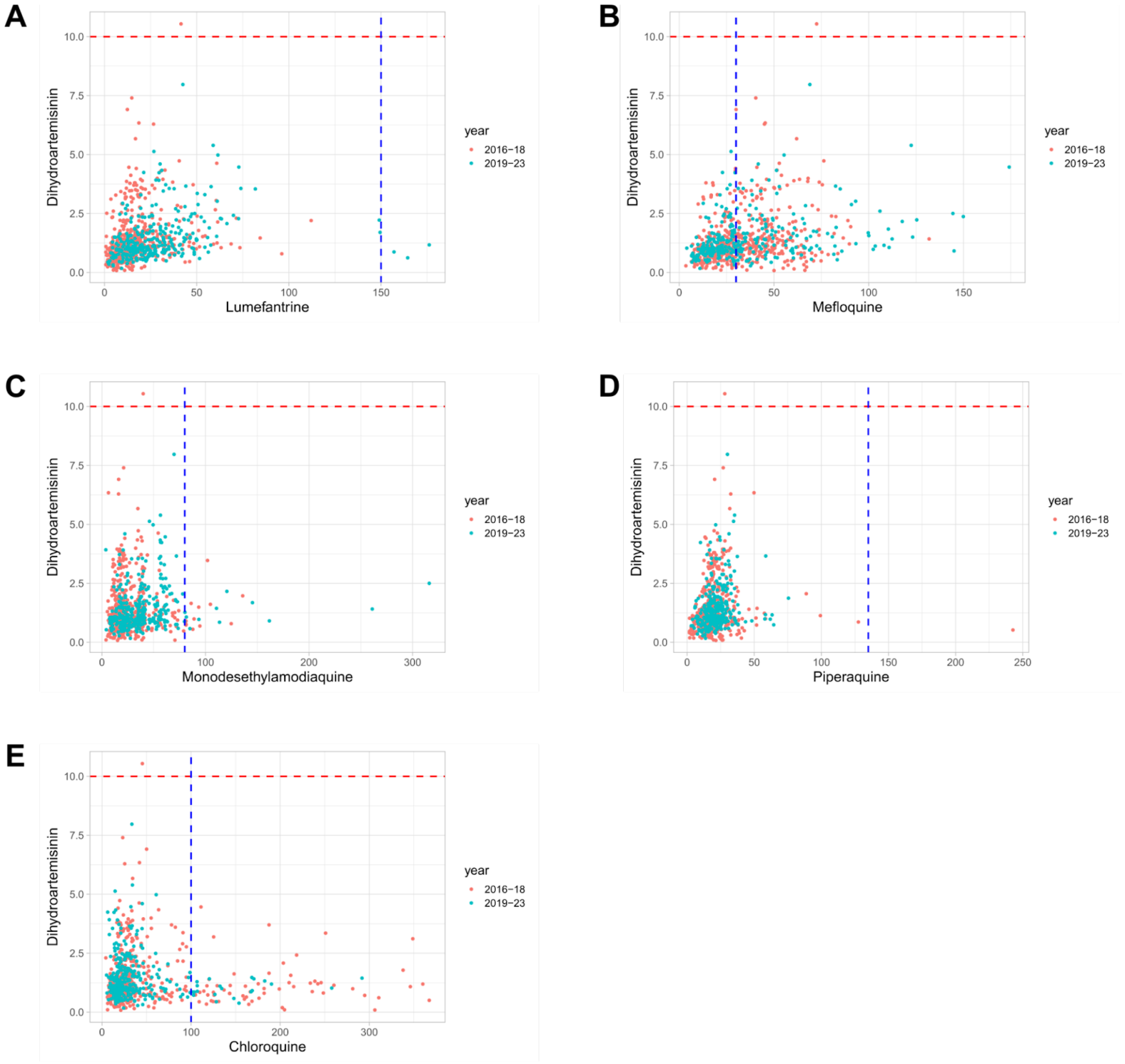


**Figure S1. Scatter plot of $IC_{50}$ for Dihydroartemisinin and partner drugs indicating $IC_{50}$ cut-offs. A)** Dihydroartemisinin (10nM) vs lumefantrine (150nM). **B)** Dihydroartemisinin (10nM) vs mefloquine (30nM). **C)** Dihydroartemisinin (10nM) vs monodesethylamodiaquine (80nM). **D)** Dihydroartemisinin (10nM) vs piperaquine (135nM). **E)** Dihydroartemisinin (10nM) vs chloroquine (100nM).

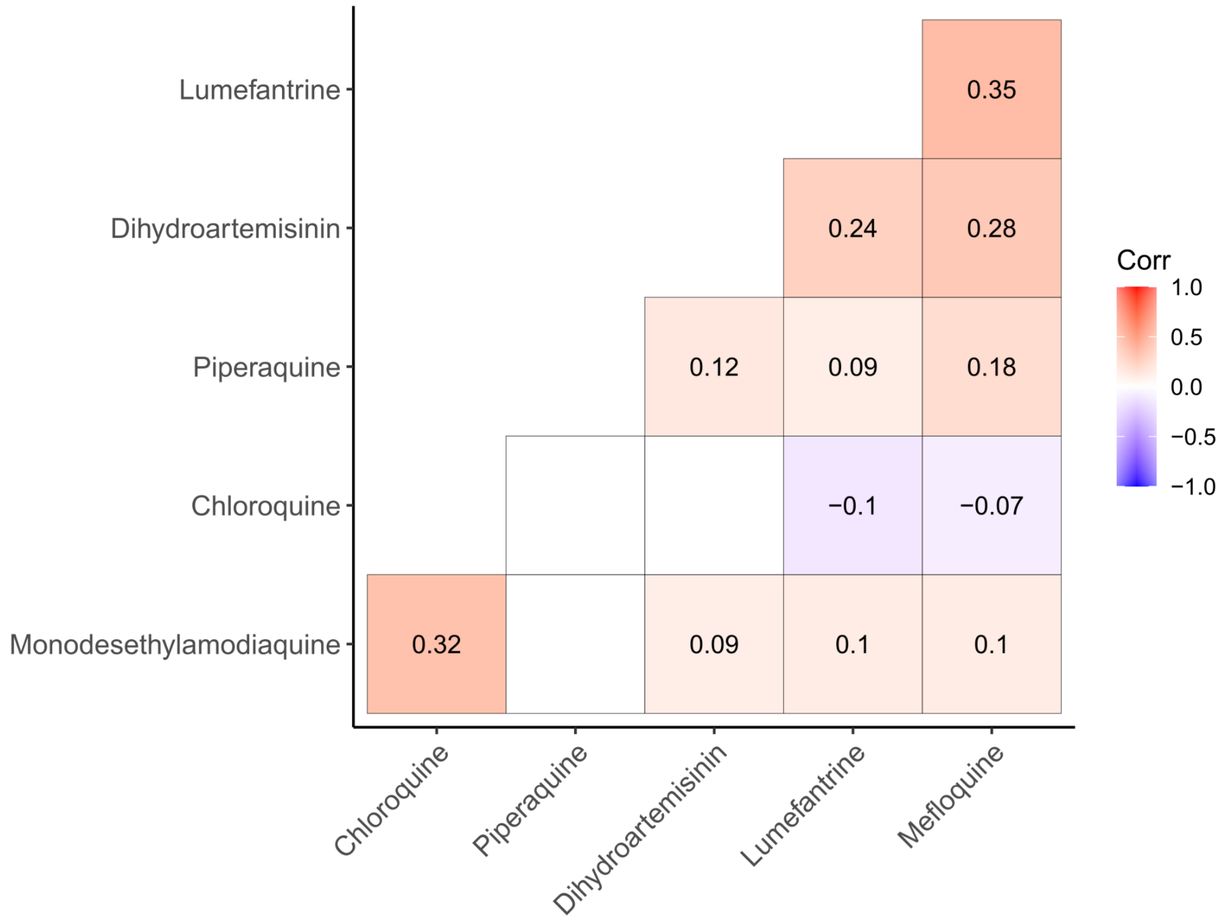


**Figure S2. Correlation between pairs of $IC_{50}$ values.** Only significant correlations are shown (r Pearson correlation: -0.07 - 0.35- $P < 0.05$).

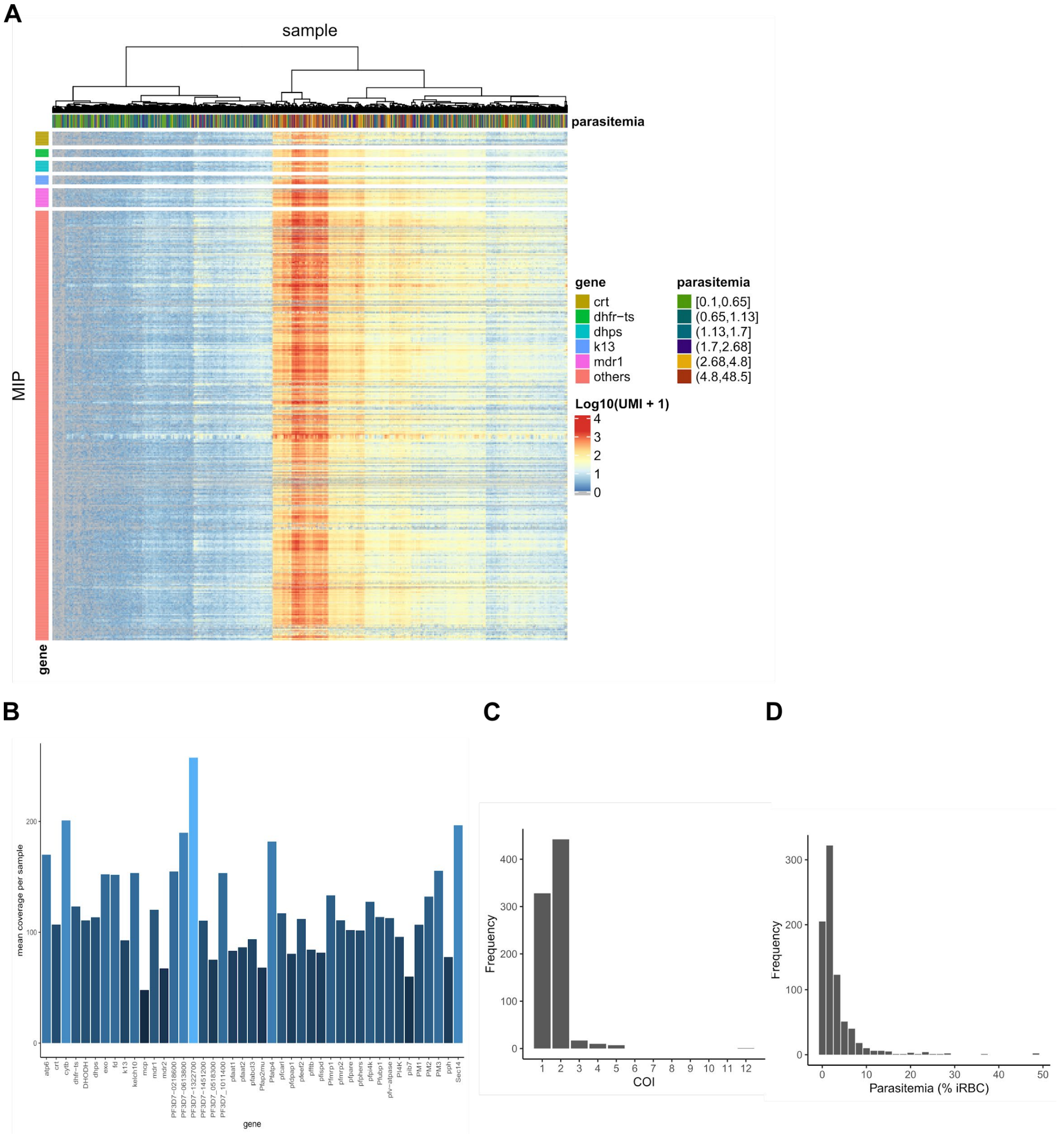


**Figure S3. MIP sequencing coverage- complexity of infections and parasitemia of isolates.** A) Heatmap showing the read counts (log10) of 810 MIPs in 805 sequenced isolates. Row annotations highlight five out of 45 genes covered by MIPs. Parasitemia of isolates is indicated by top vertical bars (% of infected red-blood-cells). B) Mean coverage per sample per gene. C) Distribution of complexity of infection (COI) in isolates. E) Distribution of parasitemia in isolates in percentage of infected red-blood-cells (iRBC).

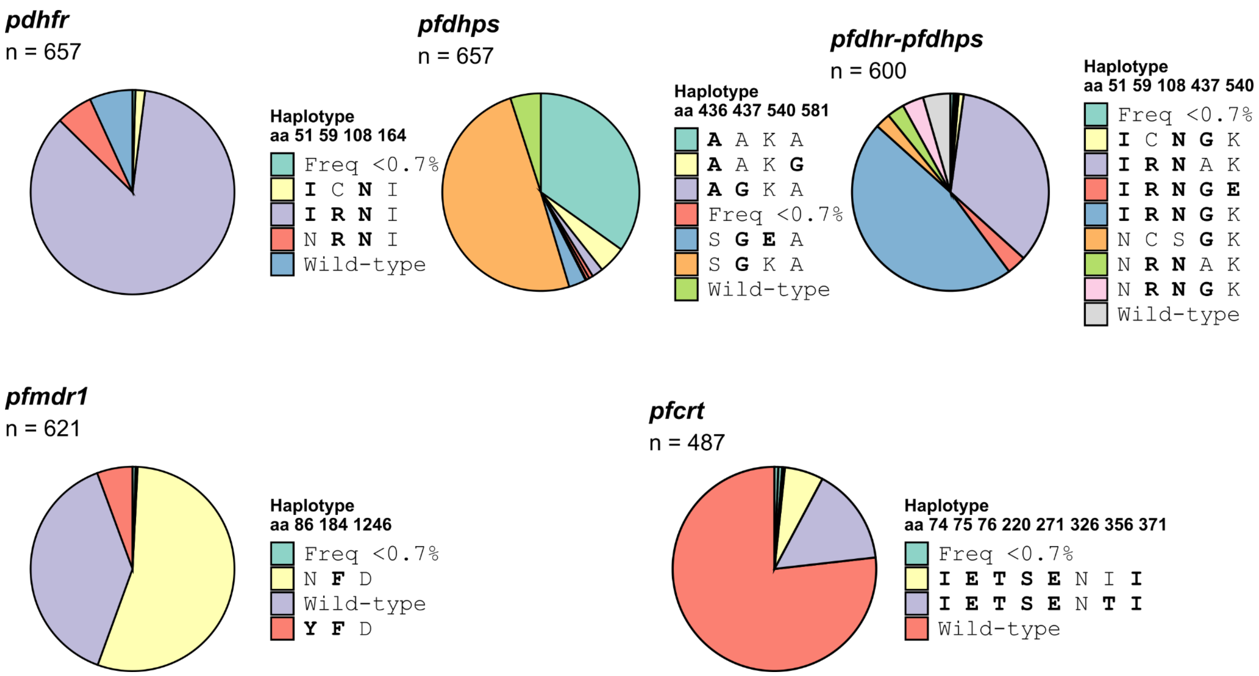


**Figure S4. Pie charts of *pfdhfr*, *pfdhps*, *pfdhfr-pfdhps*, *pfmdr1* and *pfcrt* haplotypes.** Bold letters indicate amino acid mutations whereas normal text is wildtype. Haplotypes with a frequency of less than 0.7% are not described in the graphs.

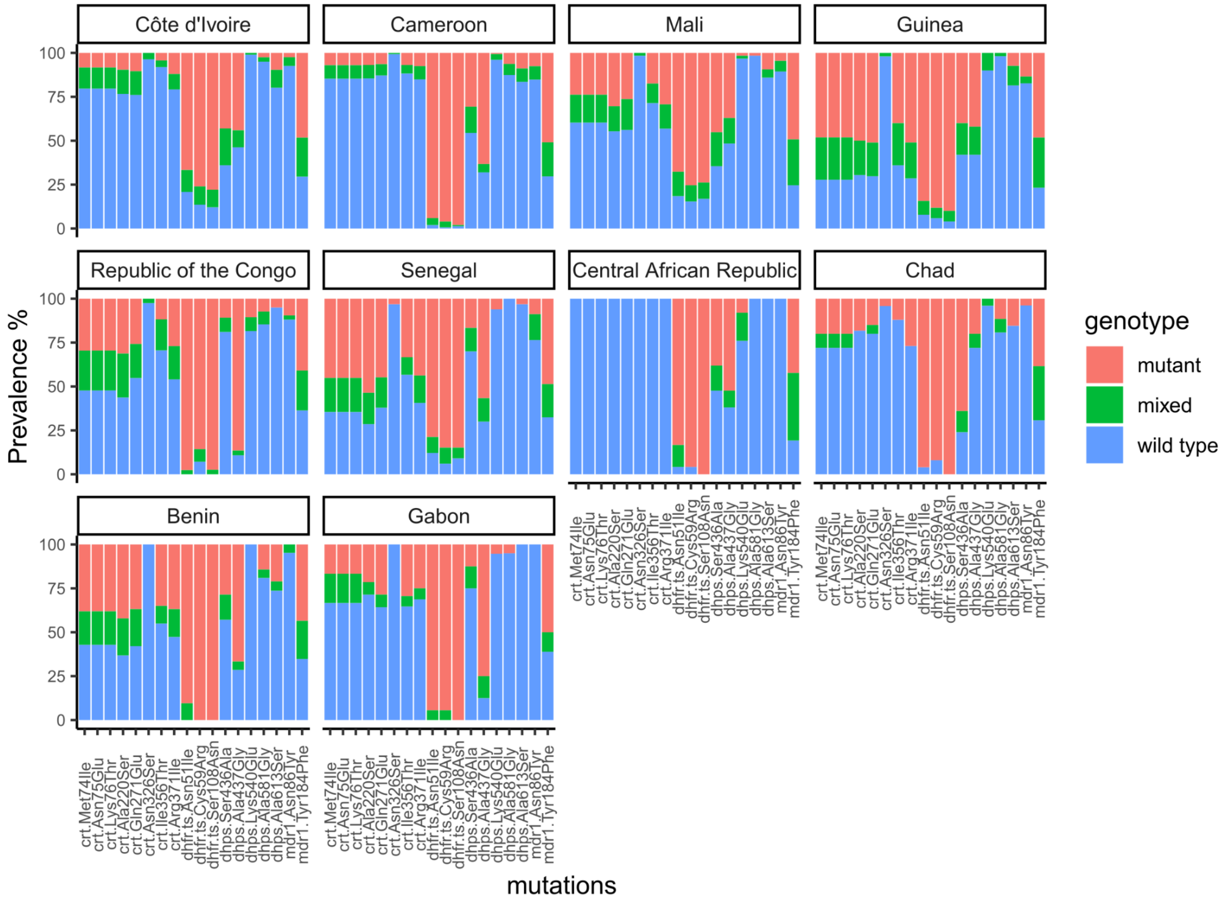


**Figure S5. Prevalence of key mutations in the ten most visited countries by participants.** Barplots display the cumulative proportion (%) of wild-type- mutant- and mixed-genotype infections per SNP.

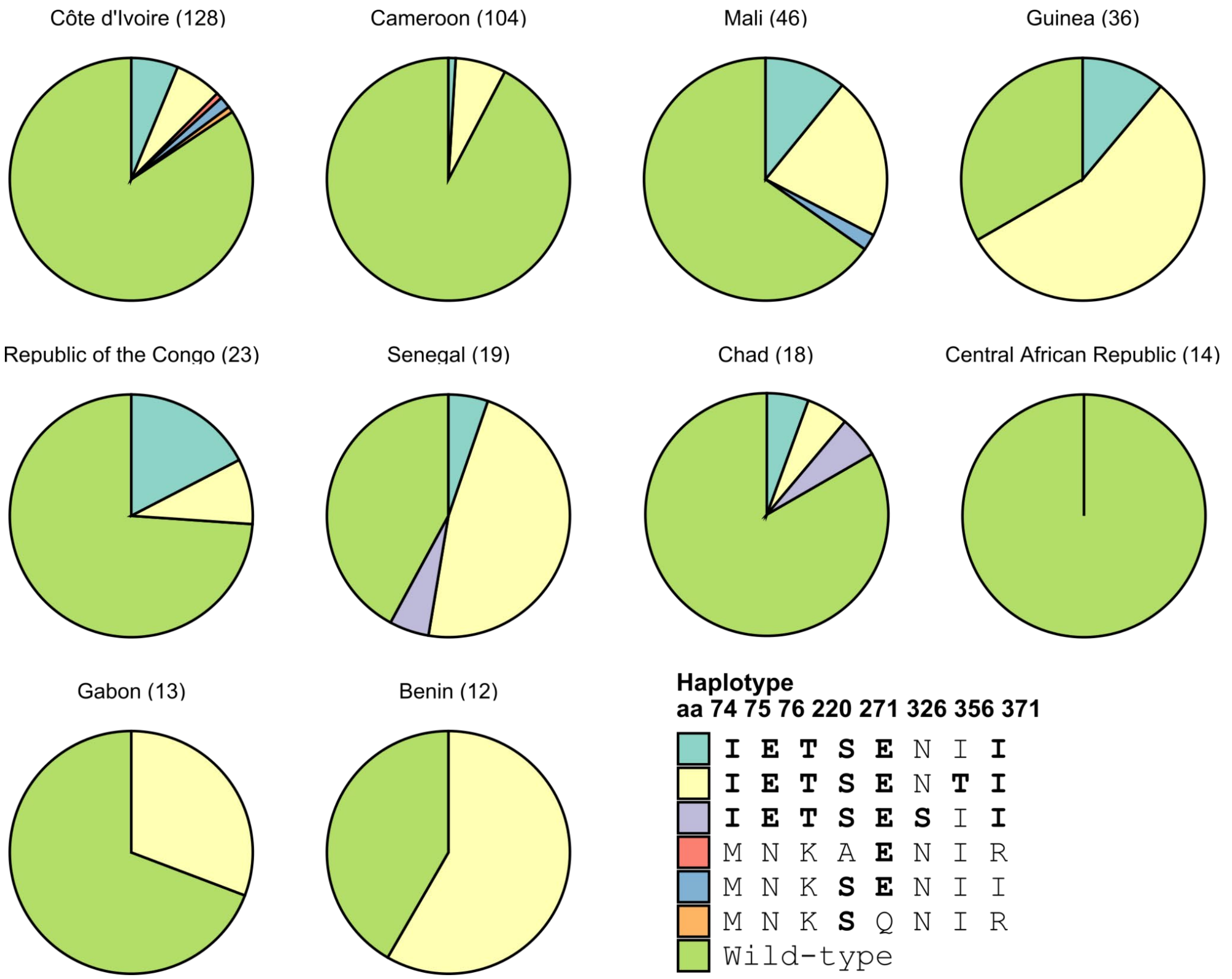


**Figure S6. Frequency distribution of the *pfcrt* haplotypes in the ten most visited countries by participants.** Letters in bold represent the amino acid changes. Haplotypes were built using the major allele per position- i.e. allele whose within sample frequency was >75 %; while alleles within sample frequency between 25% to 75% were considered unresolved. Analysis was performed on 487 DNA samples that passed this criteria. The sample size per country is indicated in parentheses.

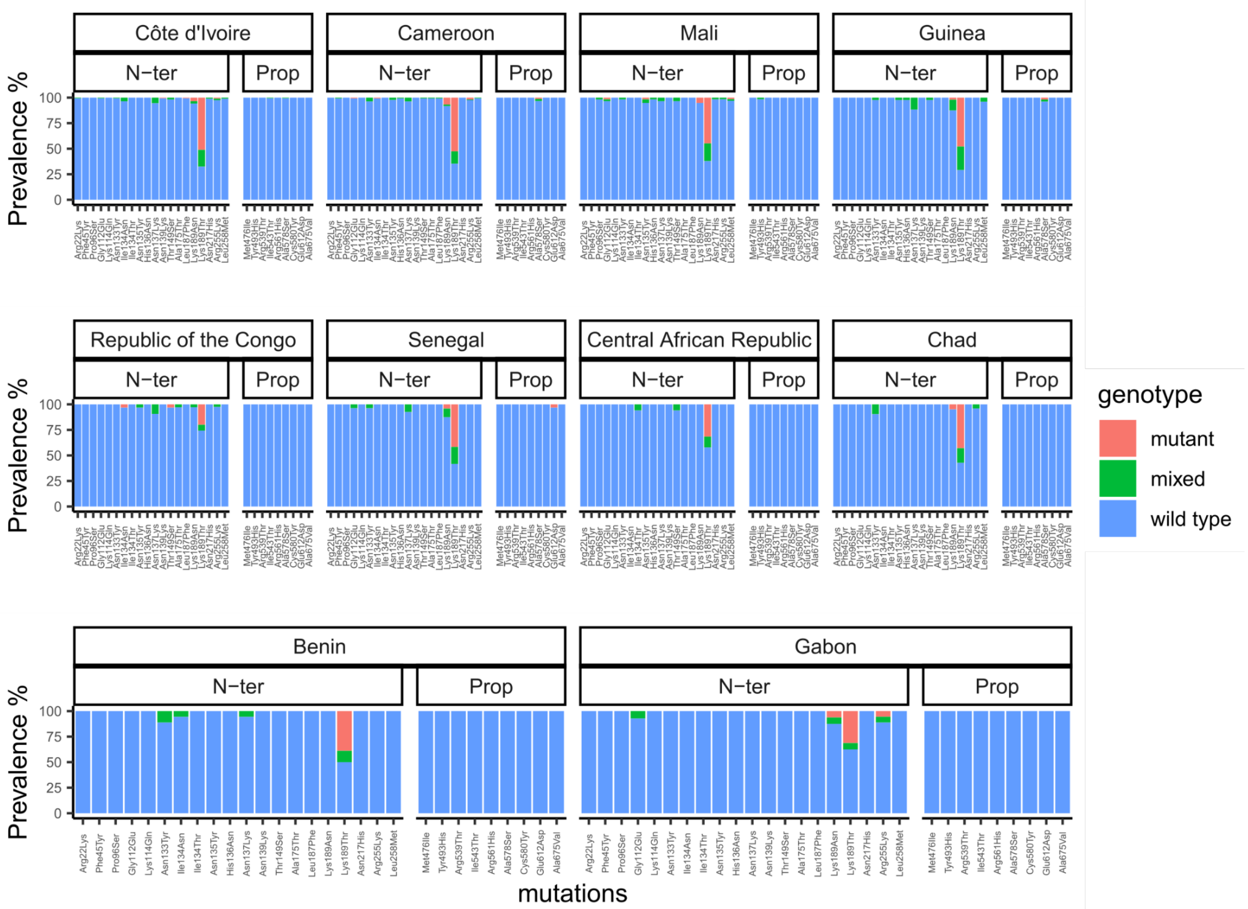


**Figure S7. Prevalence of mutations in *pfkelch13* gene in the ten most visited countries by participants.** Barplots display the cumulative proportion (%) of wild-type- mutant- and mixed-genotype infections per SNP.

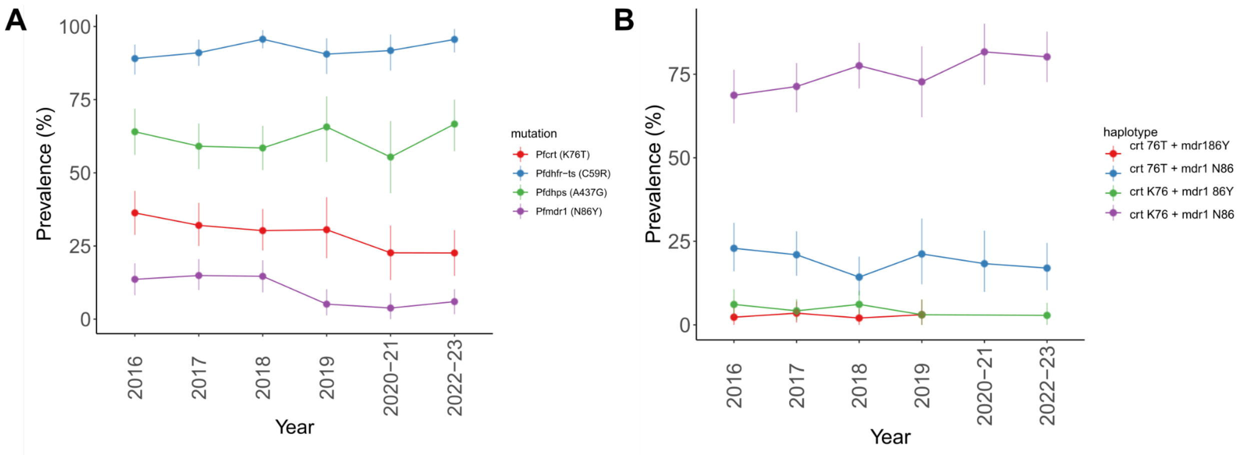


**Figure S8. Temporal change of key resistance mutations and haplotypes by year. A)** Prevalence is estimated for representative SNPs in loci: *pfcrt- pfdhfr- pfdhps* and *pfmdr1*. **B)** Prevalence of *pfcrt* 76-*pfmdr1* 86 haplotypes. 95% confidence intervals are represented by error bars.

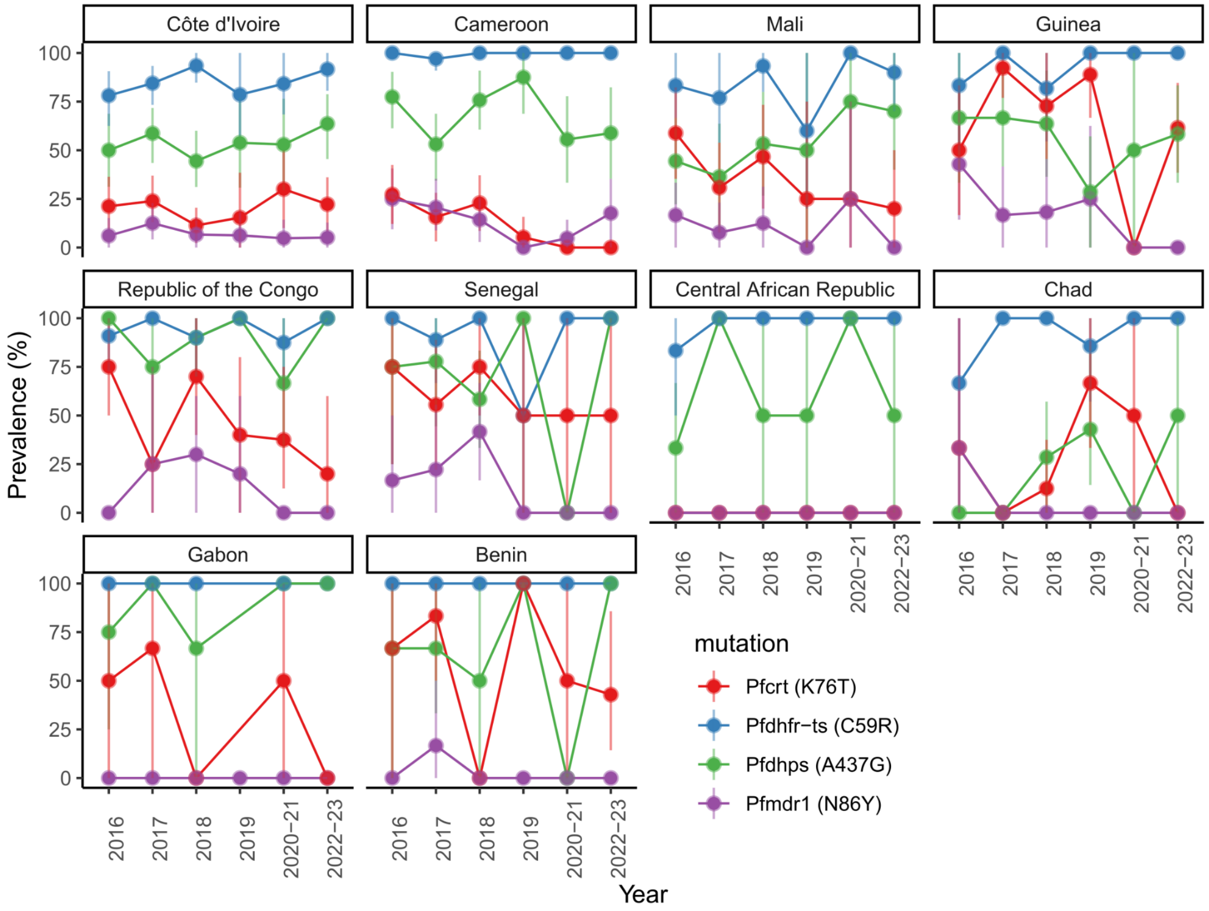


**Figure S9. Prevalence of key resistance mutations by year and by country.** Only the top ten countries were included. Prevalence is estimated for representative SNPs in loci: *pfcrt*- *pfdhfr*- *pfdhps* and *pfmdr1*. 95% confidence intervals are represented by error bars.

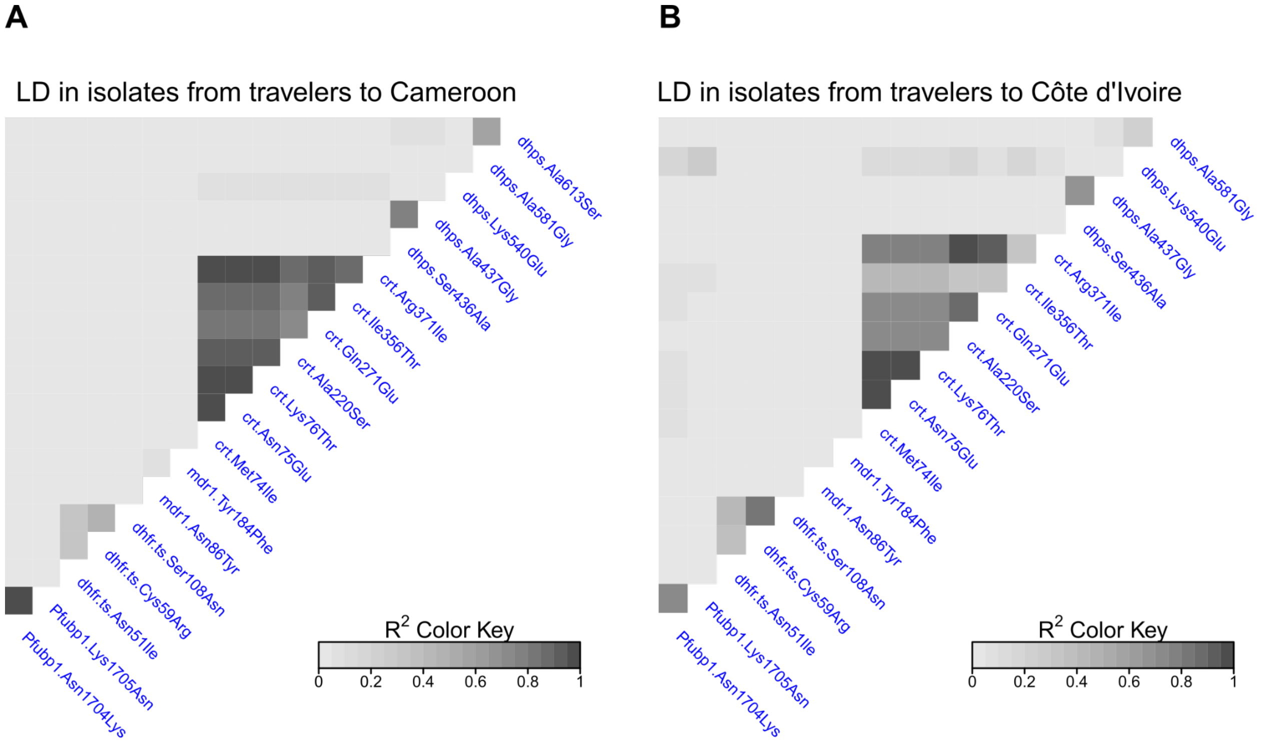


**Figure S10. Linkage disequilibrium of SNPs associated with drug resistance in isolates from travelers to Cameroon (A) and Ivory Coast (B).** Correlations between alleles are displayed in a heatmap. Biallelic SNPs with MAF > 0.01 and within-sample majority alleles were considered for this analysis. Mixed genotype infections with allele frequencies that did not exceed the threshold of 75% were discarded. LD: Linkage disequilibrium

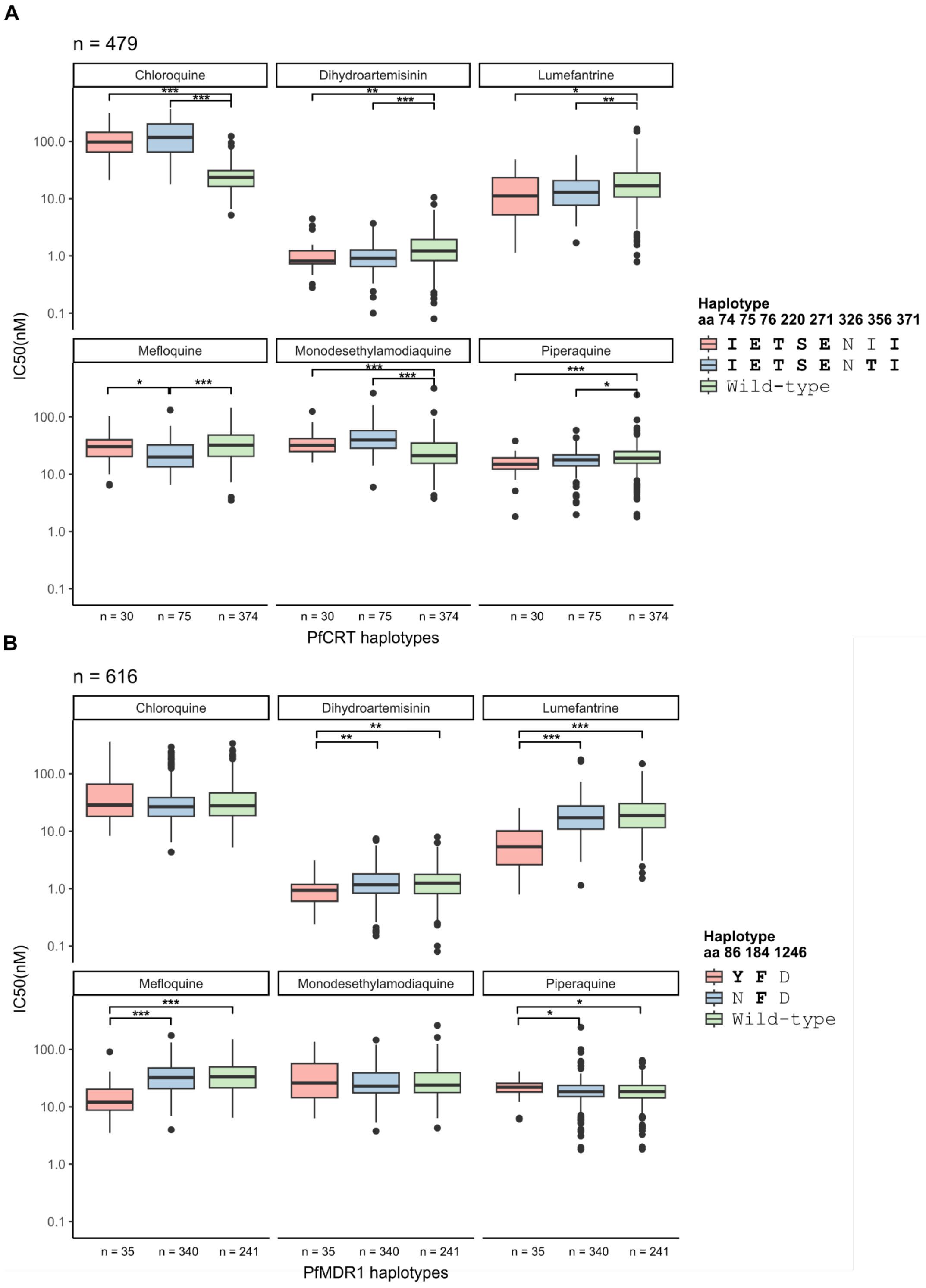


**Figure S11. Effect of *pfcrt* and *pfmdr1* haplotypes on the $IC_{50}$ of the six drugs. A)** *pfcrt* haplotypes built with amino acid positions 74- 75- 76- 220- 271- 326- 356 and 371. Haplotypes with frequency of less than 1% are not shown. Letters in bold represent the amino acid mutations. B) *pfmdr1* haplotypes built with amino acid positions 86- 184- and 1246. Haplotypes were built using the major allele per position- i.e. allele whose within sample frequency was >75 %; while alleles within sample frequency between 25% to 75% were considered unresolved. Pairwise Wilcoxon tests with Benjamini-Hochberg correction. *: $p<0.05$; **: $p<0.01$; ***: $p<0.001$.
